# Estimating the risk of post-COVID condition (PCC) in deprived communities, migrants and ethnic minorities in England: Findings from Virus Watch - a prospective community cohort study

**DOI:** 10.1101/2024.11.26.24317965

**Authors:** Wing Lam Erica Fong, Sarah Beale, Vincent Grigori Nguyen, Jana Kovar, Alexei Yavlinsky, Andrew C Hayward, Ibrahim Abubakar, Sander MJ van Kuijk, Robert W Aldridge

## Abstract

**Background:** Deprived communities, migrant and ethnic minorities were disproportionately affected by COVID-19 and may, therefore, be at a higher risk of post-COVID condition (PCC). This analysis, using data from the Virus Watch study, investigates how deprivation, migration status, and ethnic minority status influence PCC risk in both the full cohort (all regardless of infection status) and those with a confirmed COVID-19 infection.

**Methods:** A subset of participants from Virus Watch, a prospective community cohort study in England, were included. We used logistic regression to compare the predicted probability of developing PCC in both full and infected cohorts among different deprivation levels, migration and ethnic minority status categories by sex-at-birth during pre-Omicron and Omicron periods, adjusting for socio-demographic covariates.

**Results:** During the pre-Omicron period, PCC probability increased with deprivation levels, especially in females (most deprived: 7.8%, 95% CI 4.6-11.0%; least deprived: 3.5%, 2.5-4.5%). Migrant and ethnic minority males had a higher likelihood of PCC than their respective counterparts, particularly in the full cohort for migrants (6.3%, 1.8-10.8%) and the previously infected cohort for ethnic minorities (38.8%, 21.2-56.4%). However, these disparities were less pronounced in females. In the Omicron period, these differential probabilities were also less evident.

**Conclusion:** Our findings suggest that greater PCC probability among these populations is driven by increased infection risk and post-infection determinants. This underscores the need for policies and interventions to reduce infection risk and affordable and easily available healthcare services for those with PCC.

**What is already known on this topic:**

- Deprived, migrant, and ethnic minority groups experienced disproportionate rates of SARS-CoV-2 infection, hospitalisation and death due to a range of socioeconomic factors.
- Emerging evidence suggests that individuals living in deprived areas may have a higher risk of developing post COVID condition (PCC), while mixed results were reported in research on PCC risk in ethnic minorities.
- There is limited evidence on the association between migration status and PCC.

**What this study adds:**

- During the pre-Omicron period, more deprived populations were associated with increased PCC probability, especially among females. Migrant, ethnic minority and migrant-ethnic minority males also had higher PCC probability compared to their counterparts after accounting for potential confounders.
- In the Omicron period, the disparities in the likelihood of PCC were less pronounced, though migrant-ethnic minority females still had higher PCC probability, especially in those with confirmed infections, suggesting post-infection determinants play a role.
- Our study suggests pre- and post-infection determinants of PCC among these populations.

**How this study might affect research, practice or policy:**

- This study underscores the need to address health inequalities faced by deprived, migrant and ethnic minority populations to reduce their risk of COVID-19 infection and PCC.
- Further research into the intersection of migration, ethnicity, and deprivation, along with other factors such as sex and occupation, is required to guide targeted public health efforts and resource allocation.
- Target policies should address occupational and housing exposure risks, increase vaccine uptake, provide culturally appropriate public health information and ensure equitable access to PCC and primary care services

## Introduction

Migrants, ethnic minorities and individuals in deprived areas have historically experienced health inequalities which were exacerbated by the coronavirus-2019 (COVID-19) pandemic(1). The pandemic also led to the emergence of post-COVID condition (PCC) or long COVID, a syndrome comprising a wide range of symptoms which emerge within three months of acute infection and persist for at least two months with no other explanation. Globally, 10-30% of non-hospitalised and 50-70% of hospitalised cases reported developing PCC(2).

Known PCC risk factors include increasing age and female sex, and severe COVID-19 illness, but the role of socioeconomic factors remains unclear (2). Deprived communities, migrants and ethnic minorities experienced higher rates of SARS-CoV-2 infection, hospitalisation and death compared to the general population, particularly during the Wild-type, Alpha and Delta variants waves (3–5). Socio-economic disparities, including household overcrowding, occupational exposures, and limited healthcare and vaccine access, contributed to these increased risks (6–9). These communities also have higher rates of underlying health conditions, such as diabetes and hypertension, which increase their vulnerability to severe COVID-19 (10). As a result, they may be at greater risk of developing PCC.

Emerging evidence suggests that individuals in deprived areas may be at elevated risk of developing PCC. Multiple UK studies have consistently reported greater odds/risk of PCC in the most deprived decile/quintile compared to the least deprived after adjusting for covariates

(11). However, studies on PCC risk in ethnic minorities yielded mixed results. Research from the UK, US, Netherlands and Denmark (12–16) observed higher PCC odds/risk among ethnic minorities compared to the majority ethnic population. In contrast, others reported no association (17,18) or lower PCC odds/risk (19,20). Evidence on migration status is limited, but a study in the Netherlands found higher PCC risk in migrant hospitalised patients compared to non-migrants following confounder adjustment (13).

Given these inconsistencies and gaps in research, we aimed to use data from the Virus Watch prospective community cohort study in England (21) to investigate how PCC risk varies by deprivation levels, migration and ethnic minority status during pre-Omicron and Omicron periods. We evaluated this in the full cohort, irrespective of infection status (objective 1), and in individuals with confirmed COVID-19 infection (objective 2). This will provide evidence for policies addressing health inequities related to PCC and future pandemics.

## Methods

### Ethics Approval and Consent

Virus Watch was approved by the Hampstead NHS Health Research Authority Ethics Committee: 20/HRA/2320, and conformed to the ethical standards set out in the Declaration of Helsinki. Participants provided informed consent for all aspects of the study.

### Study design and participants

Virus Watch is a large prospective community cohort study established in June 2020 on the transmission and burden of COVID-19 in England and Wales (n = 58,497 as of March 2022). Participation was self-selected and limited to households with internet access and a lead household member proficient in English (Consent forms and participant information sheets were available in multiple languages). Participants complete weekly online questionnaires regarding COVID-19 symptoms, testing and vaccination, and occasional in-depth surveys on related topics, such as long-term symptom monitoring. The full study design and methodology have been described elsewhere (22).

Participants in this analysis were a subset of the Virus Watch study cohort. Inclusion criteria were: 1) registered with an English postcode, 2) responded to at least one survey on new persistent symptoms between February 2020 and March 2023, and 3) if infected, able to assign the infection variant based on SARS-CoV-2 test reporting date.

### Exposures

We examined three exposures: Index of Multiple Deprivation (IMD) as a measure of deprivation, migration status, and minority ethnicity status.

IMD was categorised into quintiles (1st quintile = most deprived, 5th = least deprived), and was based on self-reported postcode of residence linked to the May 2020 ONS Postcode Lookup (23). The 5th quintile was used as the reference category.

Migration status was determined by self-reported country of birth. Non-UK-born participants were classified as migrants, while those with missing migration status were analysed as a separate category. Comparisons used the UK-born group as the reference category.

Ethnic minority status was based on self-reported ethnicity. Participants who reported to be white Irish, white Other, mixed, South Asian, other Asian, Black, or other were categorised as ethnic minorities. The white British group acted as the reference category.

### Outcome

PCC (yes/no) was phenotypically defined using the World Health Organisation consensus definition as the onset of at least one symptom lasting at least two months within three months of acute SARS-CoV-2 infection, unexplained by other diagnoses. SARS-CoV-2 infections before the end of nationally funded COVID-19 testing (31s March 2022) were identified using linked test results from the national testing database, self-reported/study-based COVID-19 and antibody test results (see ‘Sources of SARS-CoV-2 Infection’ in Supplemental materials).

Persistent symptom tracking was based on four occasional surveys on long-term symptoms (February 2021, May 2021, March 2022, and March 2023), where participants reported any symptoms lasting at least four weeks, the onset dates of their three most severe symptoms, and whether they were ongoing or resolved. Data were available from February 2020 to March 2023, with symptom onset dates matched to track symptom episodes across multiple surveys.

PCC cases were defined as individuals with a confirmed SARS-CoV-2 infection within three months of symptom onset with at least one symptom lasting two months. Symptom duration was calculated from symptom onset to survey completion (for ongoing symptoms) or reported resolution date. Only PCC cases reported by 30 June 2022 (three months post nationally-funded testing end) were included to minimise misclassification. Participants who were hospitalised for/with COVID-19 or reported ongoing symptoms lasting less than two months were also excluded. Non-PCC participants completed all four surveys and either reported no persistent symptoms or symptoms without a confirmed SARS-CoV-2 infection within three months of onset, reducing the risk of undetected or unreported symptoms.

### Covariates

The covariates considered were age group (<25, 26-44, 45-64, 65+ years), sex at birth (male and female) and occupation. Occupations were categorised into five categories based on occupation type, exposure risk, and employment status: higher exposure risk occupation, lower exposure risk occupation, not in employment, retired, and unknown/other status (see Supplemental Table 1 and further details elsewhere) (24).

SARS-CoV-2 infection variants were based on regional variant dominance periods (25,26). Study periods were categorised as ‘pre-Omicron’ (1 February 2020 - 13 December 2021) and ‘Omicron’ (14 December 2021 - 12 June 2022) (Supplemental Table 2).

### Statistical analyses

We summarised baseline sociodemographic and clinical characteristics using descriptive statistics.

Separate analyses were conducted for each exposure using two distinct cohorts. The first included all participants, regardless of SARS-CoV-2 infection history, to assess the association between each exposure and PCC. The second included individuals with a confirmed SARS-CoV-2 infection to estimate the association between each exposure and the risk of PCC within infected populations, providing an estimate of post-infection PCC probability differences among different exposure groups.

Directed acyclic graphs were used to identify minimally sufficient adjustment sets for each exposure (Supplemental Figures 1-3). For IMD quintile, the adjustment set included age, migration status, ethnic minority status, and occupation for objective 1, and age, migration and ethnic minority status for objective 2. For migration status, the set included age and minority ethnicity status for both objectives, while for minority ethnicity status, the adjustment set consisted of age.

We conducted stratified multivariable logistic regression analyses for each exposure and cohort, stratified by sex and variant period. To explore the intersection of minority ethnicity and migration status, we performed an additional analysis using a composite variable that combined these two factors, adjusted for age (27). Furthermore, a sensitivity analysis was conducted by categorising white British participants with missing migration status as UK-born, based on evidence from the 2011 Census that 98% of white British individuals were UK-born (28). Multicollinearity was tested (see ‘Multicollinearity’ in Supplemental Materials). Results were expressed as predicted probabilities (PP) to allow comparison across all exposures, and odds ratios (OR) to assess the relative odds of developing PCC in each exposure group compared to the reference group in both cohorts.

## Results

Figure 1 illustrates participant selection. Table 1 reports the sociodemographic and clinical characteristics of the analysis cohorts. See Supplemental Tables 6-8 for characteristics by each exposure.

**Figure 1.**
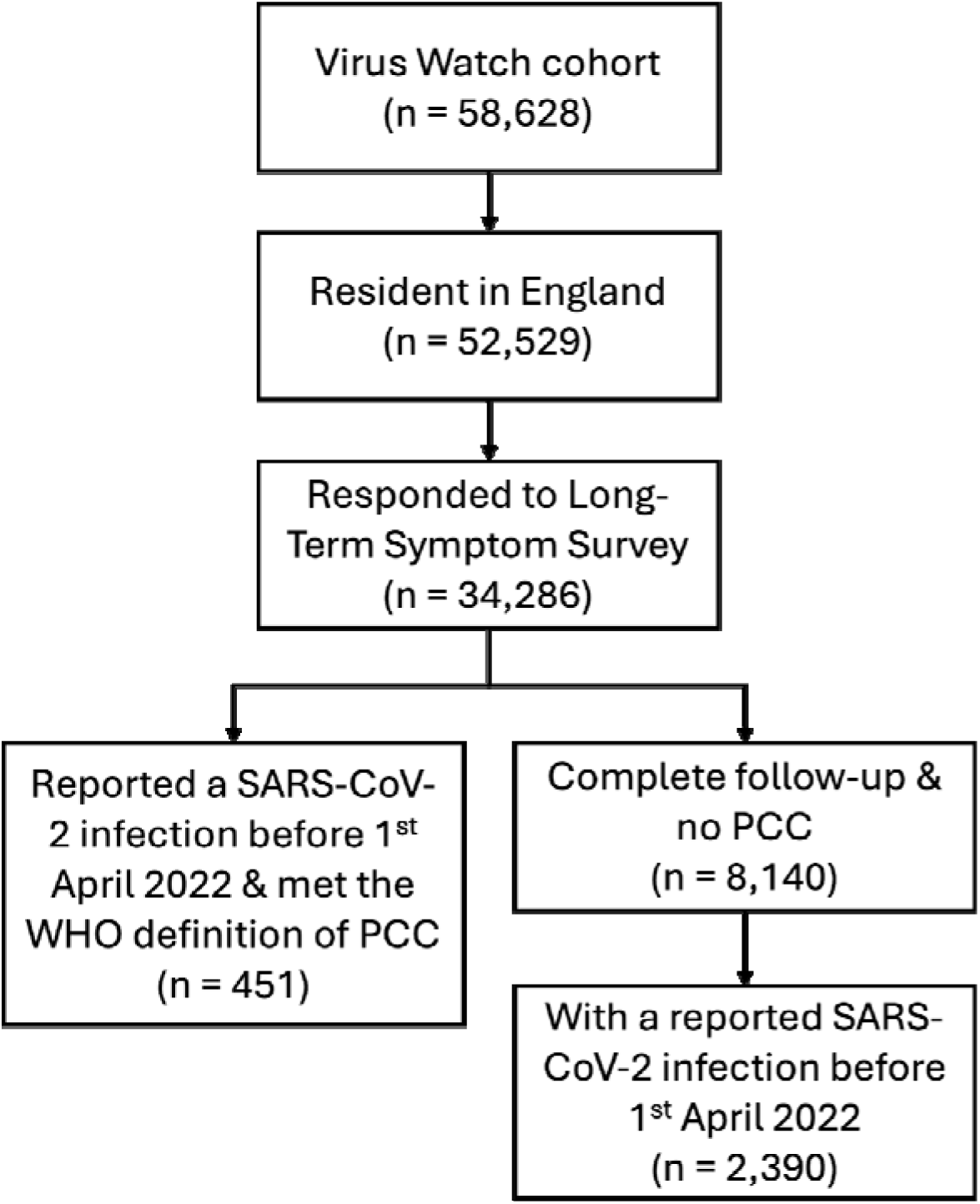
Flow diagram of participant eligibility.

**Table 1.** Demographic and clinical characteristics of the analysis cohorts.

| Characteristic | All participants<br>regardless of any confirmed<br>SARS-CoV-2 infection<br>N = 8,591 | Participants with a reported<br>confirmed SARS-CoV-2<br>infection<br>N = 2,841 |
| --- | --- | --- |
| **Age group** |  |  |
| 0-24 | 415 (4.8%) | 231 (8.1%) |
| 25-44 | 477 (5.6%) | 267 (9.4%) |
| 45-64 | 3,185 (37%) | 1,141 (40%) |
| 65+ | 4,514 (52%) | 1,202 (42%) |
| **Sex (at birth)** |  |  |
| Male | 4,906 (57%) | 1,655 (58%) |
| Female | 3,685 (43%) | 1,186 (42%) |
| **Ethnic minority status** |  |  |
| White British | 7,870 (92%) | 2,578 (91%) |
| Minority ethnic | 666 (7.8%) | 250 (8.8%) |
| Missing/Prefer not to say | 55 (0.6%) | 13 (0.5%) |
| **Migration status** |  |  |
| UK Born | 7,299 (85%) | 2,355 (83%) |
| Not UK Born | 537 (6.3%) | 171 (6.0%) |
| Missing | 755 (8.8%) | 315 (11%) |
| **IMD Quintile** |  |  |
| 1 (most deprived) | 593 (6.9%) | 213 (7.5%) |
| 2 | 1,139 (13%) | 406 (14%) |
| 3 | 1,734 (20%) | 573 (20%) |
| 4 | 2,274 (26%) | 714 (25%) |
| 5 (least deprived) | 2,778 (32%) | 921 (32%) |
| Missing | 73 (0.8%) | 14 (0.5%) |
| **Clinical vulnerability** |  |  |
| Not clinically vulnerable | 4,856 (57%) | 1,679 (59%) |
| Clinically vulnerable | 2,391 (28%) | 763 (27%) |
| Clinically extremely vulnerable | 1,262 (15%) | 370 (13%) |
| Missing | 82 (1.0%) | 29 (1.0%) |
| **Body Mass Index (BMI)** |  |  |
| Underweight | 82 (1.0%) | 24 (0.8%) |
| Normal | 2,836 (33%) | 898 (32%) |
| Pre-obesity | 2,535 (30%) | 808 (28%) |
| Obesity | 1,361 (16%) | 459 (16%) |
| Severe obesity | 174 (2.0%) | 48 (1.7%) |
| **Missing** | 1,603 (19%) | 604 (21%) |
| **Occupation** |  |  |
| Higher Risk Occupation | 1,386 (16%) | 591 (21%) |
| Lower Risk Occupation | 1,746 (20%) | 643 (23%) |
| Not in Employment | 358 (4.2%) | 118 (4.2%) |
| Retired | 4,120 (48%) | 1,105 (39%) |
| Unknown or Other Status | 981 (11%) | 384 (14%) |

Across all exposure groups, the likelihood of developing PCC was higher during the pre-Omicron than the Omicron period, regardless of sex and infection status. Overall, the PCC probability was higher in females than males across all exposure groups (Figures 2 and 3; Supplemental Tables 9-12, 29, 30).

**Figure 2.**
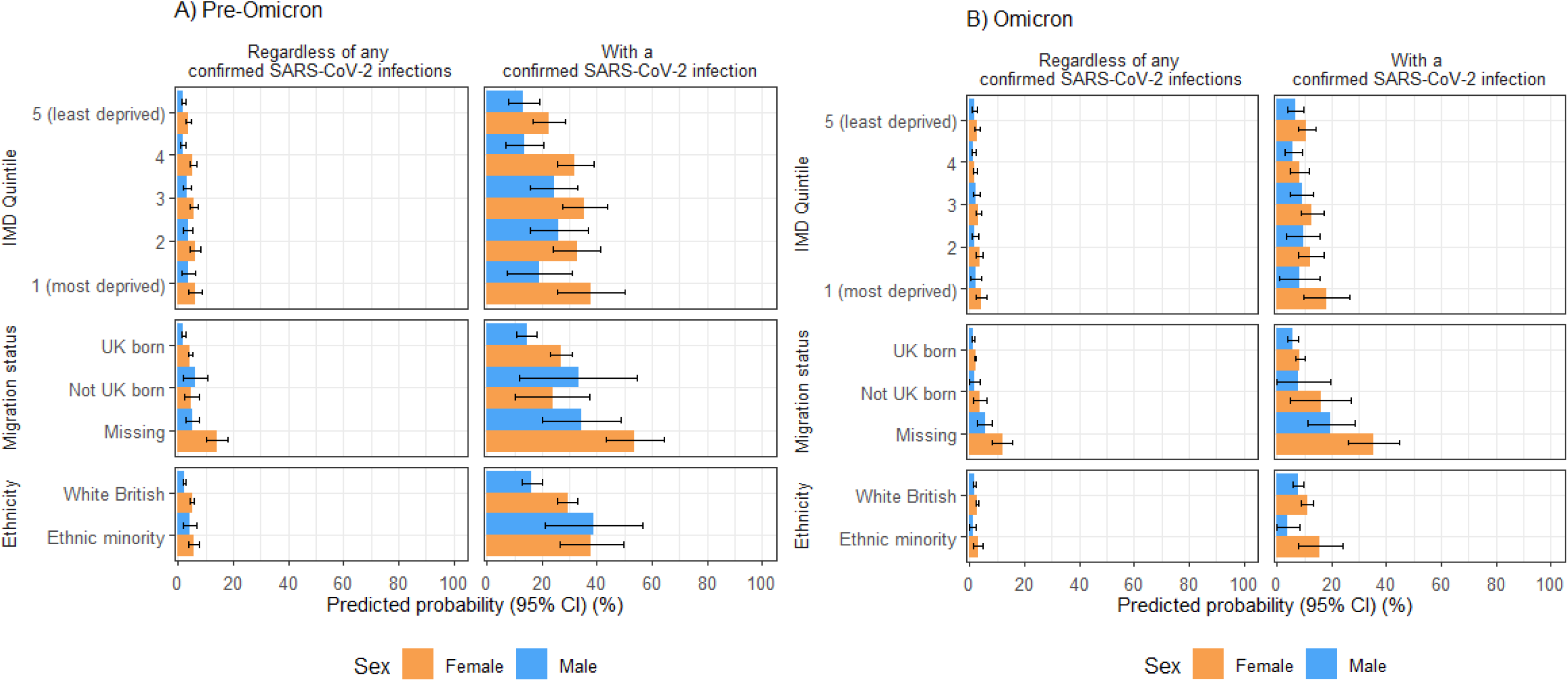
Predicted probabilities of PCC by IMD quintile, migration status and ethnic minority status in the full cohort and individuals with a previous SARS-CoV-2 infection, categorised by reported sex-at-birth during A) pre-Omicron and B) Omicron periods.

**Figure 3.**
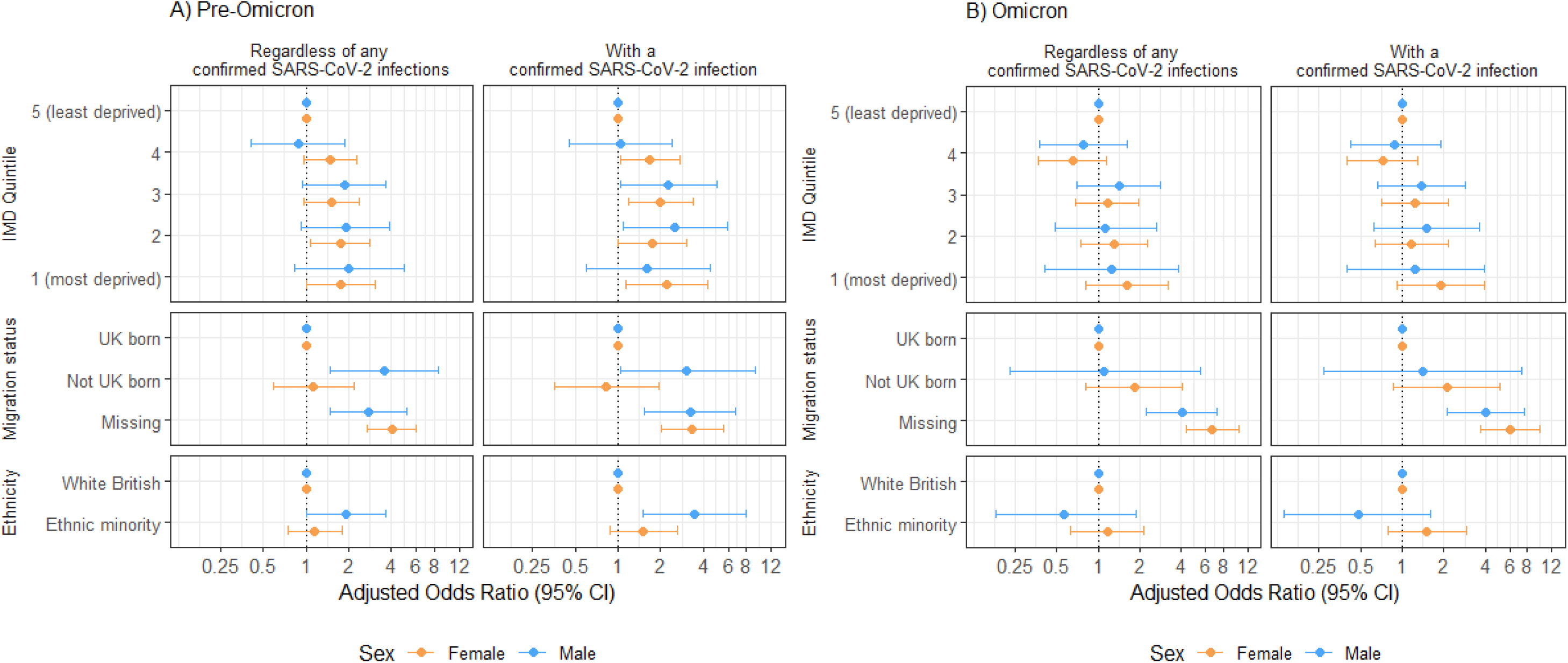
Conditional adjusted odds ratio of PCC by IMD quintile, migration status and ethnic minority status in the full cohort and individuals with a previous SARS-CoV-2 infection, categorised by reported sex-at-birth during A) pre-Omicron and B) Omicron periods.

### PCC Risk by IMD Quintile

Pre-Omicron, increasing PCC probability across deprivation levels was observed for both sexes, regardless of infection status (Figure 2A). Among individuals with confirmed infection, females residing in the most deprived quintile and males in the 2nd most deprived quintile had the highest PCC probabilities (Female: 37.7%, 95% confidence interval (CI) = 25.4-50.0%; Male: 26.1%, 15.5-36.7%) (Supplemental Table 11). There was strong evidence for an increased odds of PCC in females in the most deprived areas (adjusted OR (aOR): 2.1, 1.1-4.3) and males in the 2nd most deprived areas (2.5, 1.1-5.8) compared to counterparts in the least deprived areas (Figure 3A; Supplemental Tables 13-14).

In the Omicron period, there were no between-quintile differences for either sex in both cohorts (Figure 2B). Females in the most deprived areas had the highest PCC probability (18.1%, 9.6-26.6%), exceeding those in less deprived areas and all males (Supplemental Table 12). However, there was little evidence of increased odds in more deprived groups than those least deprived in both cohorts during this period (Figure 3B; Supplemental Tables 15-16).

### PCC Risk by Migration Status

Pre-Omicron, migrant males had a higher PCC probability compared to UK-born males in the full cohort (migrant: 6.3%, 1.8-10.8%; UK-born: 1.9%, 1.4-2.4%) and the confirmed infection cohort (migrant: 33.1%, 11.8-54.5%; UK-born: 14.4%, 10.5-18.3%) (Figure 2A; Supplemental Table 11). There was strong evidence for a 3.6 (1.5-8.5) increased odds of PCC in migrant males compared to UK-born males in the full cohort, but this was attenuated in the infected group (aOR: 3.1, 1.0-9.2) (Figure 3A; Supplemental Tables 17-18). Individuals with missing migration status had the highest PCC probabilities in both cohorts.

During the Omicron period, PCC probabilities were higher for non-UK-born than UK-born individuals in both cohorts, but no differences in PCC risk were identified, as suggested by overlapping 95% CIs were wide (Figure 2B; Supplemental Table 12). There was also little evidence to suggest differential odds between the two groups (Figure 3B). Similar to the pre-Omicron period, individuals with unknown migration status exhibited the highest PCC probability, particularly among females (female: 35.1%, 25.8-44.5%; male: 19.8%, 11.2-28.4%) (Supplemental Tables 19-20).

Reclassifying white British participants with missing migration status as UK-born attenuated the increased PCC probability seen in non-UK-born males in both cohorts during the pre-Omicron period. The increased PCC risk among those with unknown migration status was also lost (Supplemental Figure 4 & Tables 25-28).

### PCC Risk by Ethnic Minority Status

Pre-Omicron, ethnic minority males had a higher PCC probability than white British males in the full cohort, with ethnic minority males experiencing a 1.93 (1.01-3.68) higher odds of PCC. A similar trend was seen in the infected cohort (white British: 38.8%, 21.2-56.4%; ethnic minority: 16.2%, 12.5-20.0%), with the association observed being more pronounced (aOR: 3.44, 1.49-7.94). Among females, we found no evidence of an association between ethnic minority status and PCC in both cohorts (Figures 2A & 3A; Supplemental Tables 11, 21-22).

In the Omicron period, PCC probability was generally low across all groups, regardless of infection status. Among infected individuals, the likelihood of PCC was higher in ethnic minority than white British females (white British: 11.0%, 9.0-13.0%; ethnic minority: 15.7%, 7.5-23.8%) but lower in ethnic minority males than white British males (white British: 7.8%, 5.8-9.7%; ethnic minority: 3.9%, 0.0-8.3%), though confidence intervals overlapped (Figure 2B; Supplemental Table 12). Adjusted ORs suggested no differences in the odds of developing PCC between ethnic groups for either cohort and sex (Figure 3B; Supplemental Tables 23-24).

When including the ‘ethnicity-migration status’ composite variable, among the infected cohort, we found strong evidence for a higher PCC probability in male ethnic minority migrants (49.7%, 26.4-73.1%) compared to white British UK-born males (13.7%, 9.9-17.4%) during the pre-Omicron period, and in female ethnic minority migrants (19.4%, 6.9-31.8%) than their white British UK-born counterparts (8.2%, 6.4-10.1%) during the Omicron period (Figure 4; Supplemental Tables 29-34).

**Figure 4.**
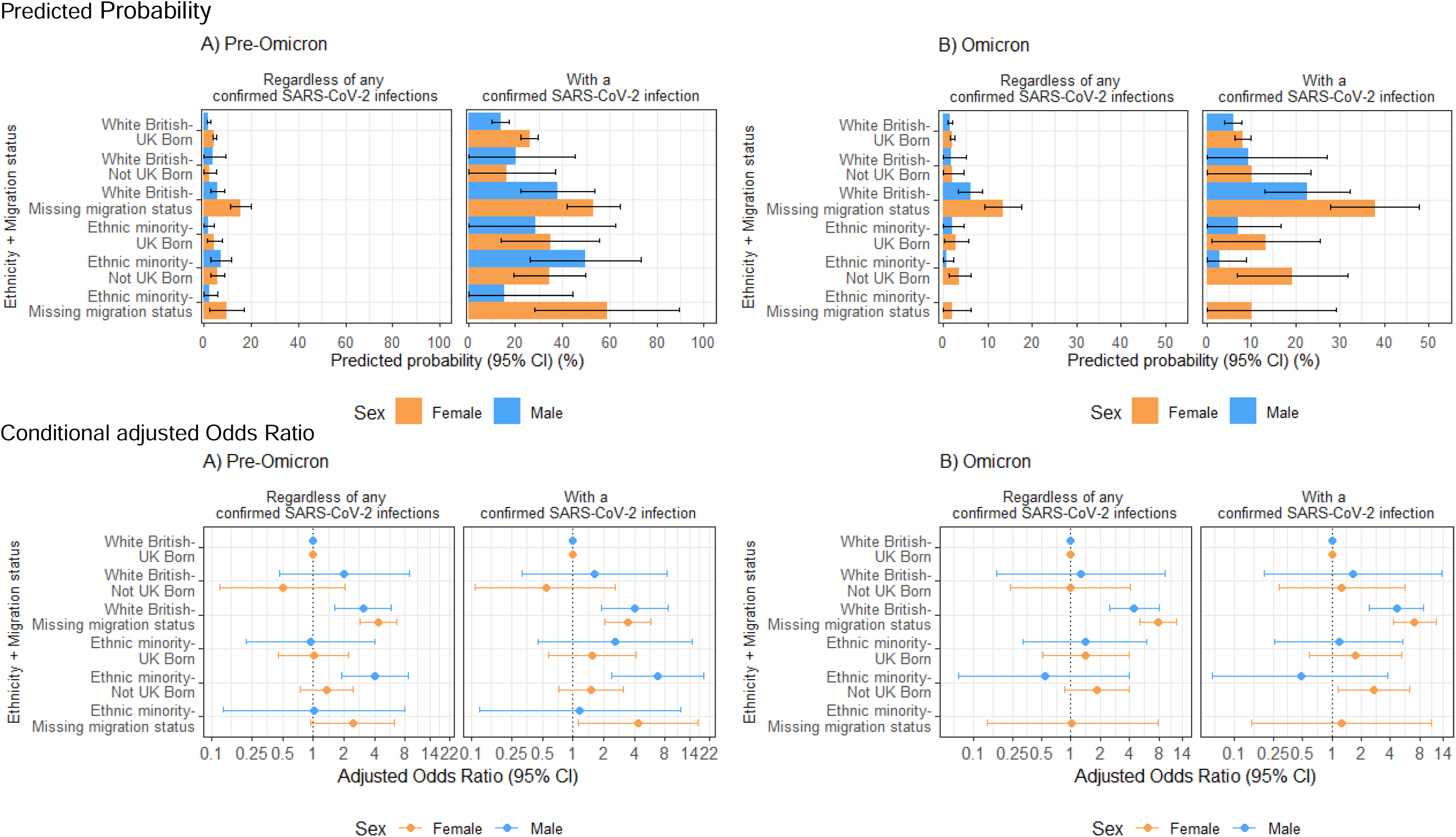
Predicted probability and conditional adjusted odds ratio of PCC by the ‘ethnicity-migration’ composite variable in the full cohort and individuals with a previous SARS-CoV-2 infection, stratified by reported sex-at-birth during A) pre-Omicron and B) Omicron periods. No PP and OR estimates are available for males in the ‘ethnic minority-missing migration status’ group from the Omicron period due to the small sample size.

## Discussion

Using data from a community cohort in England, we found that deprivation levels were associated with higher PCC probability, particularly among females, during the pre-Omicron period. Migrant and ethnic minority groups had higher PCC probabilities, though these disparities were less pronounced in females - who overall had a higher probability of PCC - and more pronounced in migrant-ethnic minority males. These disparities were also less pronounced in the Omicron period, with some evidence for an association with PCC for migrant-ethnic minority females than their white British-UK-born counterparts.

While our study lacked statistical power to demonstrate an association between deprivation and PCC risk, the point estimates suggested one, which is consistent with broader literature. Shabnam et al. found higher PCC risk in females than males across all IMD deciles and a dose-response relationship with deprivation levels in both sexes (11). Our findings regarding elevated PCC probability among ethnic minority males align with some existing studies, although findings on ethnic disparities have been mixed (12–20). Furthermore, our results corroborate those from the Netherlands, which reported higher PCC risk in migrants (13). It is important to note that our findings are not directly comparable since our analyses were stratified by sex and variant period. Our findings underscore the intersectionality of sex, ethnicity, migration and deprivation as PCC determinants and the need for further comparative research to better understand these intersections.

Our findings suggest that inequalities in the likelihood of PCC were influenced by pre- and post-infection determinants. Migrant and ethnic minority males, often employed in insecure or frontline roles, reliant on public transport, working or residing in overcrowded environments, and experiencing pre-existing health inequalities, likely faced heightened exposure risk (6–9). These findings may reflect intervention-induced inequalities during the pre-Omicron period - where measures including lockdowns, social distancing and work-from-home, while lowering overall infection rates, disproportionately impacted these populations depending on work and housing-related exposure. Differential vaccine access in these populations may also affect their infection risk (29). However, by the Omicron period, as restrictions were lifted, the observed inequality gaps narrowed when infection became more widespread.

Slower vaccine uptake among migrants with non-White ethnicity, along with lower vaccination rates in ethnic minority and deprived groups, as reported by Burns et al. and Curtis et al., likely also contributed to the increased risk of infection and severe illness, particularly by pre-Omicron variants (30–32). These variants were associated with more severe COVID-19 illness, a key infection-related PCC determinant (33). Improved vaccination coverage across different populations and Omicron’s reduced pathogenicity contributed to milder illness and, subsequently, the observed attenuation of PCC risk disparities in the later period (32,34). This underscores the need to improve vaccine access and uptake in deprived and minority groups, particularly in future pandemics, to reduce their infection and PCC risk.

Delayed care for COVID-19, often due to institutional and personal barriers to healthcare access, may lead to more severe illness and increase the risk of PCC (35). The individual and intersecting characteristics of migrants and ethnic minorities also translate to unique challenges related to healthcare access, such as cultural and language differences, fear of stigma, and distrust of healthcare systems (35). Job insecurity and lack of paid sick leave, common among deprived communities and in some migrant and ethnic minority groups, also delay or prevent timely care, as many cannot afford to miss work or risk losing their jobs (35,36). These challenges are more pronounced for women in these populations as they are more likely to work in low-income, precarious jobs and serve as primary caregivers (37).

### Strengths and limitations

To our knowledge, this is the first study to examine the association between migration status and PCC in England, assess how deprivation, migration and minority ethnicity status influence PCC risk over time during pre-Omicron and Omicron periods, and explore whether PCC determinants of these populations arise before or after a SARS-CoV-2 infection. Virus Watch’s longitudinal design and detailed sociodemographic and clinical data collection across multiple pandemic periods, along with linkage to the national testing database, reduced recall and selection bias, strengthening the validity of study findings.

Several limitations should be considered. Using an area-level measure of deprivation limits individual-level conclusions, and categorising migration and ethnic minority status as binary variables, while necessary for adequate sample sizes, restricted a more in-depth analysis of PCC risks across specific migrant groups and ethnicities. Future studies should explore these differences more granularly. Deprived, migrant, and minority groups are likely underrepresented in Virus Watch due to language barriers, limited internet access, and economic circumstances (38). Misclassification of PCC cases may have arisen from limiting community testing, recall bias, and missed asymptomatic infections, particularly during the pre-Omicron (Wild Type and Alpha) periods.

## Conclusion

Our findings underscore the critical need to tackle health inequalities to minimise the infection risk and long-term consequences of COVID-19 and future pandemics, especially for deprived, migrant and ethnic minority groups. This includes addressing pre-infection determinants such as mitigating occupational and housing-related exposure risks. Targeted policies and interventions should focus on the specific needs of these groups in vaccination, recovery and rehabilitation efforts. Early efforts to increase vaccine uptake and ensure equitable access to PCC and primary care services in these communities are crucial to reducing their infection and subsequent PCC risk. Providing culturally and linguistically appropriate care and collaborating with trusted community partners is also necessary to disseminate accurate public health information about PCC. Further investigation into the intersectionality of multiple social dimensions, including sex and occupation, on PCC risk is also recommended to guide targeted public health efforts and resource allocation for the most vulnerable populations.

## Data Availability

Data from the Virus Watch cohort are available on ONS Secure Research Service. The data are available under restricted access as they contain sensitive health data. Access can be obtained by ONS Secure Research Service.

