## Supplementary materials for "Estimating the risk of post-COVID condition (PCC) in deprived communities, migrants and ethnic minorities in England: Findings from Virus Watch - a prospective community cohort study"

**Supplemental Materials**

**Sources of SARS-CoV-2 Infection**

We used the following sources to identify SARS-CoV-2 infections before the end of state-funded national COVID-19 testing (31st March 2022) among study participants:

1. Polymerase chain reaction (PCR) or lateral flow test (LFT) results from linkage to the Second Generation Surveillance System (SGSS). Linkage was conducted by NHS Digital, and the linkage variables were sent in March 2021. Participant data were linked over time and between databases using the unique personal identifier recorded at all interactions with the NHS number, full name, date of birth and home address. The linkage period for SGSS Pillar 1 (secondary care testing) encompassed data from March 2020 until August 2021 and from June 2020 until August 2023 for SGSS Pillar 2 (community testing).
2. Self-reported SARS-CoV-2 test results (PCR or LFT) received from outside the study (e.g. via the UK Test Trace and Isolate system or privately obtained) as part of the weekly illness survey.
3. PCR test results from swabs provided by Virus Watch to a subset of participants between October 2020 and May 2021 and between January 2023 and June 2023. Participants self-administered a PCR swab if they experienced fever, cough, loss or change of taste or smell/taste.
4. Anti-nucleocapsid or anti-spike antibody (prior to vaccination) serological test results provided by Virus Watch to a subset of participants. Participants attended in-clinic serological testing (September 2020-January 2021 and April 2021-July 2021) and/or conducted monthly at-home finger-prick serology (February 2021-April 2022). Anti-nucleocapsid or anti-spike antibodies presence in serology indicates prior infection. Infection date was estimated by identifying seroconversion during routine monthly testing. Further testing details are provided in the study protocol (1)

### **Supplementary Table 1. Occupation categories.**

| **Category** | **Included groups** |
| --- | --- |
| Higher exposure risk occupation | Healthcare |
|  | Indoor Trades, Process and Plant |
|  | Leisure and Personal Service |
|  | Sales and Customer Service |
|  | Social care and Community Protective Service |
|  | Teaching, Education and Childcare |
|  | Transport and Mobile Machine |
| Lower exposure risk occupation | Administrative and Secretarial |
|  | Managers, Directors and Senior Officials |
|  | Other Professional and Associate Occupations |
|  | Outdoor Trades |
| Retired | Retired |
| Not in employment | Not in Employment or Homemaker |
|  | Permanently sick or disabled |
|  | Student |
| Unknown/Other Status | Unknown/Other Status |

**Supplementary Table 2. Dates of variant periods by English National Region.** Missing dates indicate periods when no single variant accounted for >75% of infections. Data on Omicron sub-lineages were only available for England as a whole.

| **Region** | **Pre-Omicron period**  **(Wild-Type, Alpha, Delta)** | **Omicron period**  **(Omicron BA.1, Omicron BA.2)** |
| --- | --- | --- |
| East Midlands | 1 Feb 2020 - 11 Dec 2021 | 20 Dec 2021 - 12 June 2022 |
| East of England | 1 Feb 2020 - 11 Dec 2021 | 19 Dec 2021 - 12 June 2022 |
| London | 1 Feb 2020 - 07 Dec 2021 | 14 Dec 2021 - 12 June 2022 |
| North East | 1 Feb 2020 - 13 Dec 2021 | 22 Dec 2021 - 12 June 2022 |
| North West | 1 Feb 2020 - 11 Dec 2021 | 19 Dec 2021 - 12 June 2022 |
| South East | 1 Feb 2020 - 10 Dec 2021 | 19 Dec 2021 - 12 June 2022 |
| South West | 1 Feb 2020 - 12 Dec 2021 | 20 Dec 2021 - 12 June 2022 |
| West Midlands | 1 Feb 2020 - 12 Dec 2021 | 20 Dec 2021 - 12 June 2022 |
| Yorkshire and the Humber | 1 Feb 2020 - 12 Dec 2021 | 20 Dec 2021 - 12 June 2022 |

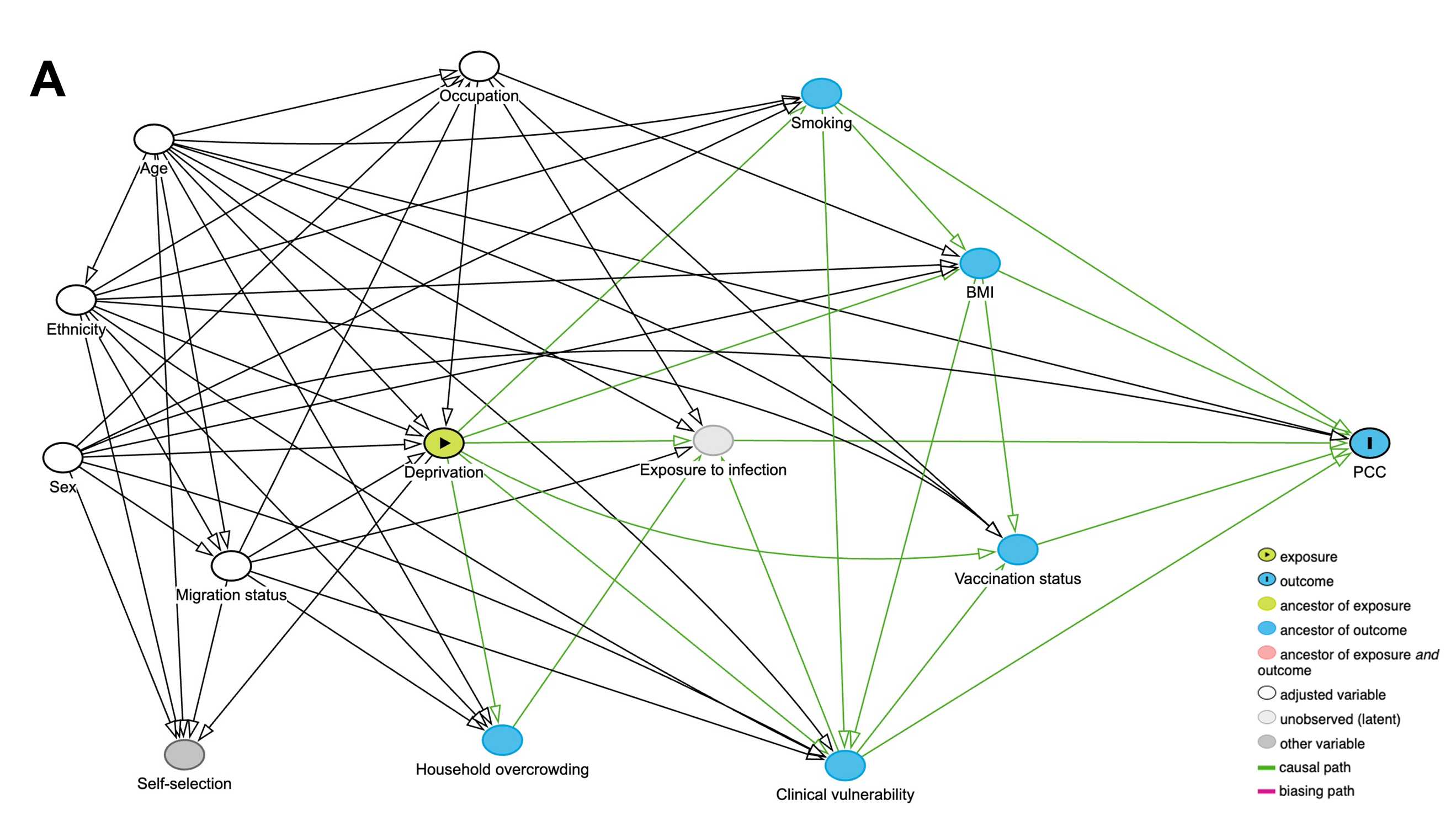

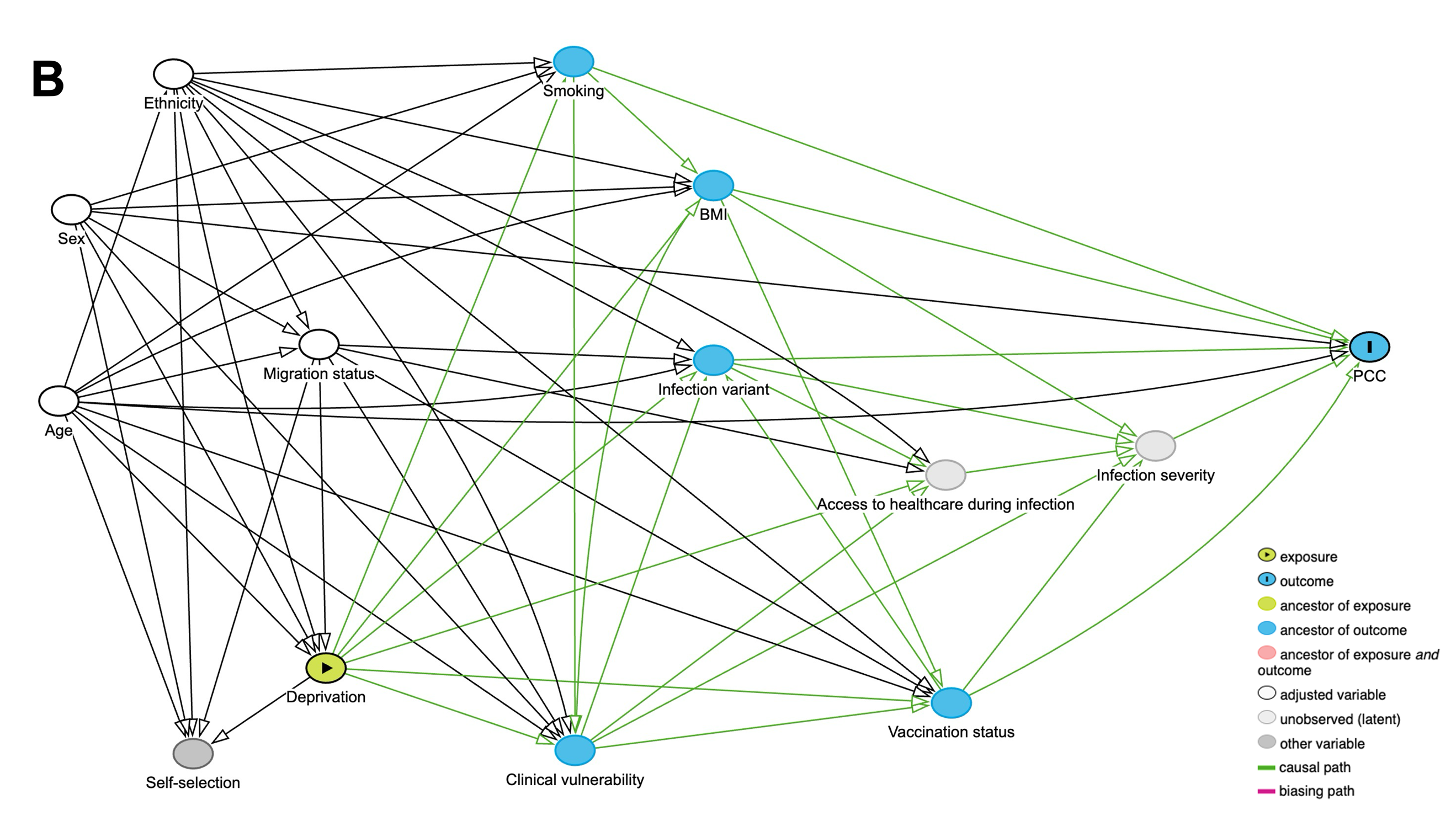

### **Supplementary Figure 1. Directed Acyclic Graph for the impact of deprivation (IMD Quintile) on the development of post-COVID condition A) regardless of any previous SARS-CoV-2 infections and B) in individuals with a confirmed SARS-CoV-2 infection.**

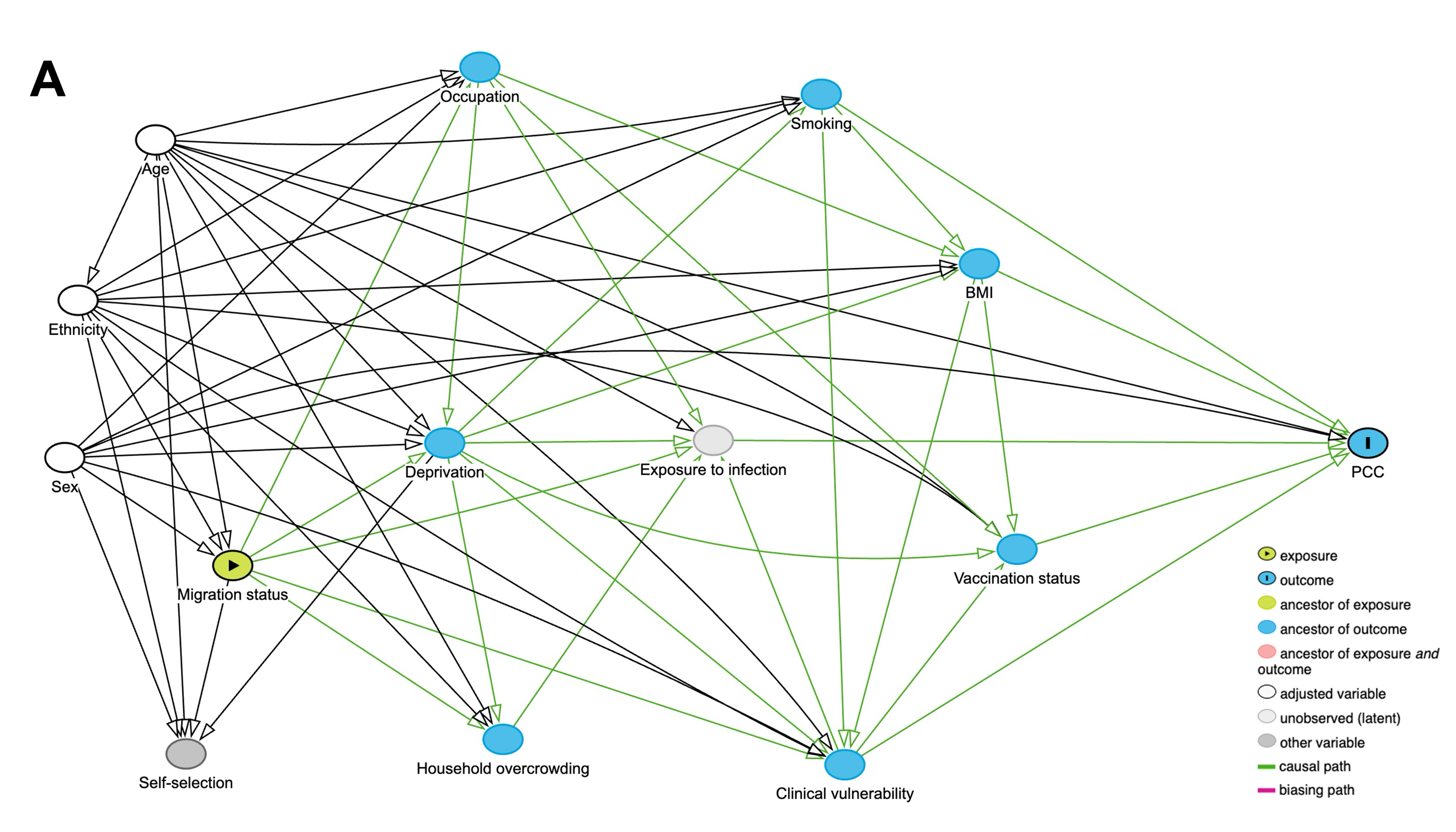

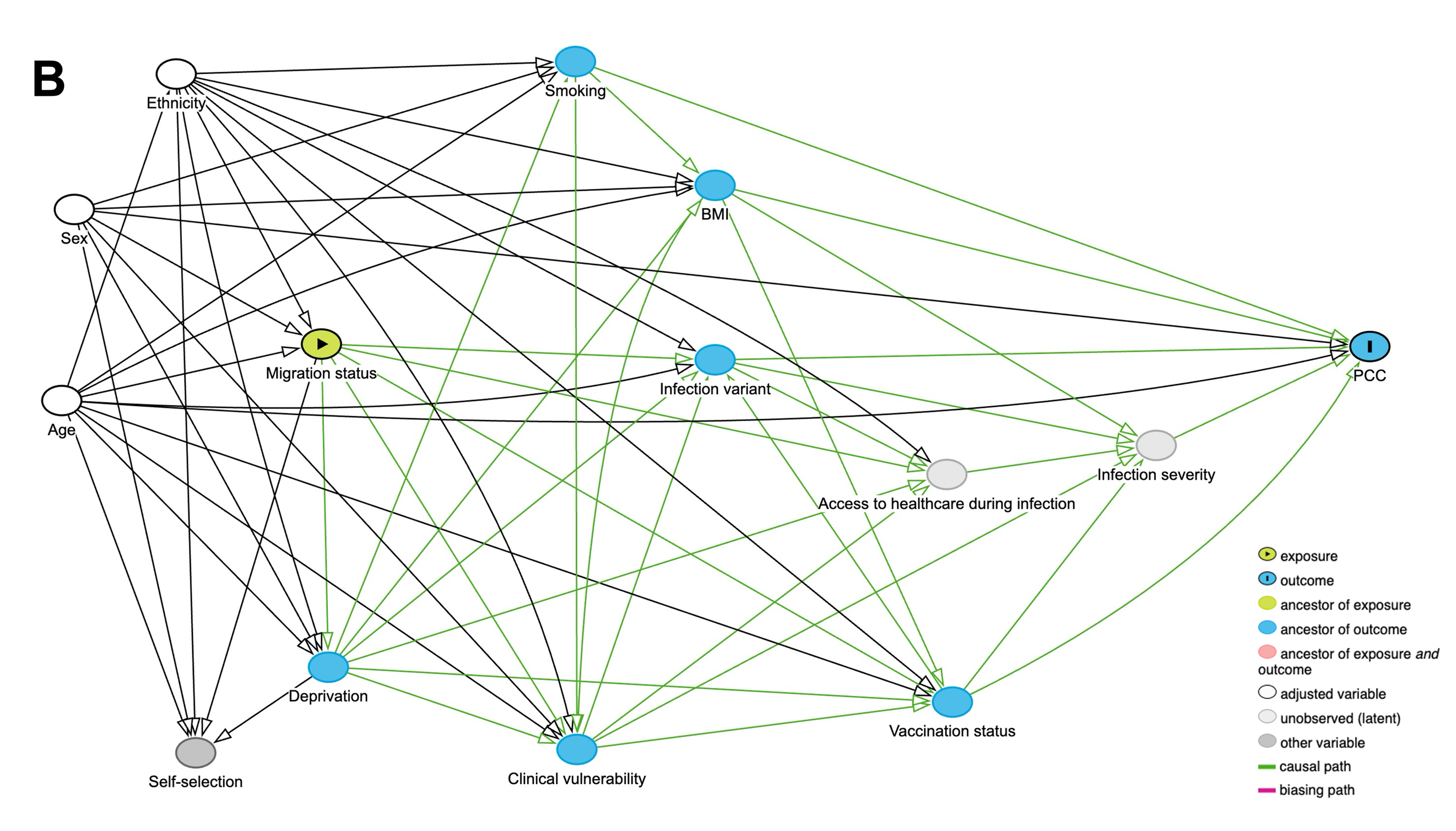

### **Supplementary Figure 2. Directed Acyclic Graph for the impact of migration status on the development of post-COVID condition A) regardless of any previous SARS-CoV-2 infections and B) in individuals with a confirmed SARS-CoV-2 infection.**

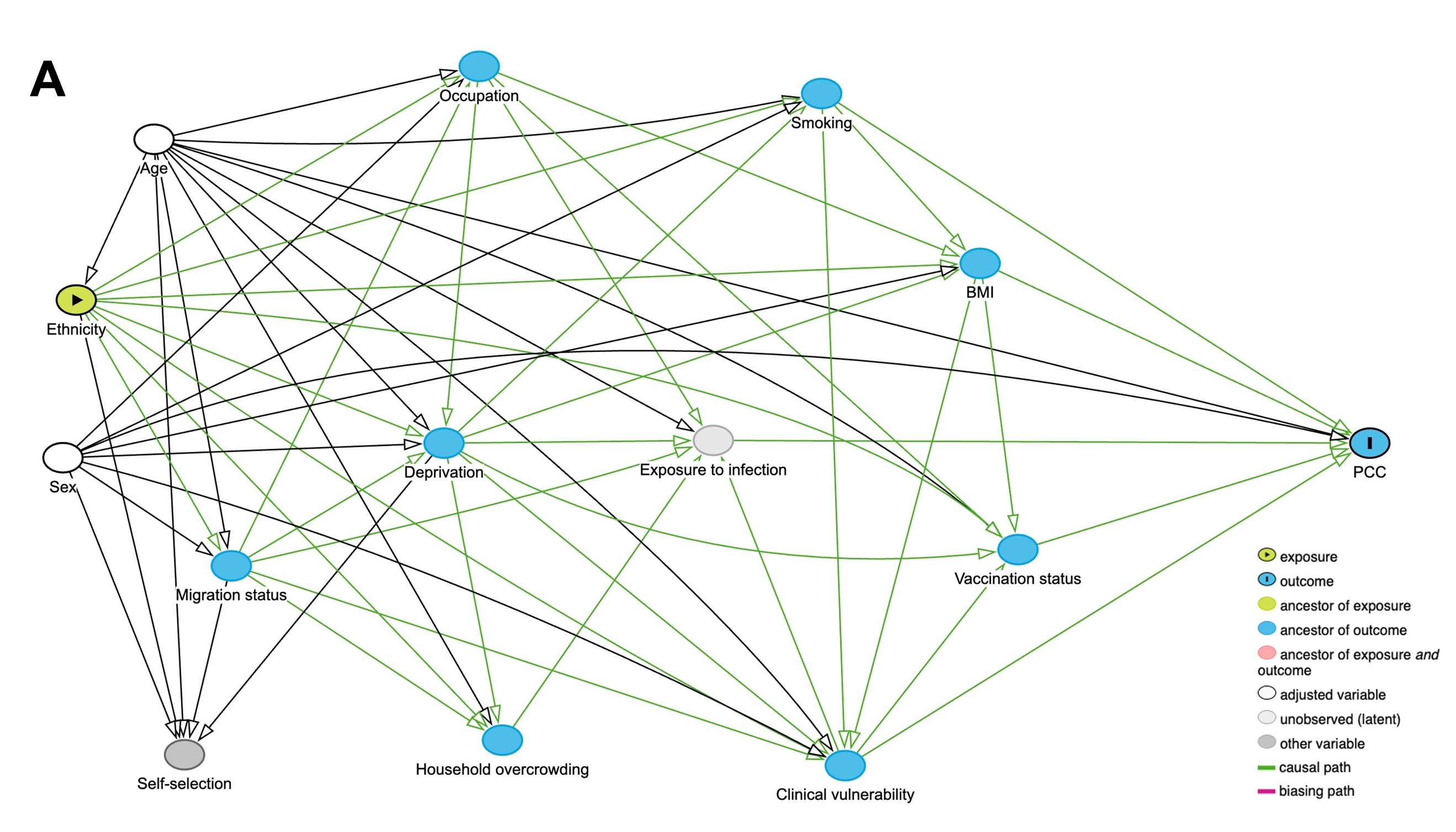

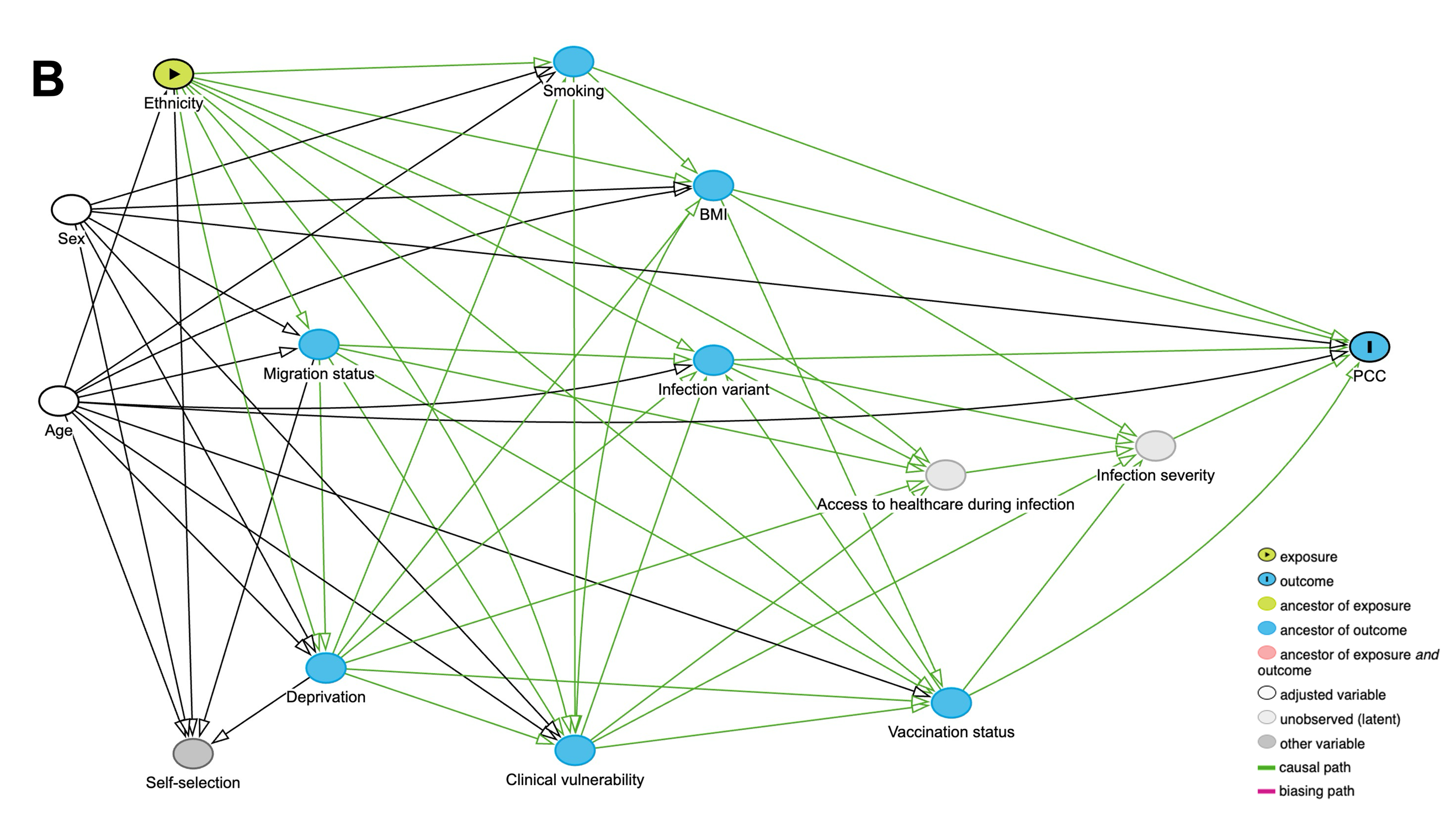

### **Supplementary Figure 3. Directed Acyclic Graph for the impact of ethnic minority status on the development of post-COVID condition A) regardless of any previous SARS-CoV-2 infections and B) in individuals with a confirmed SARS-CoV-2 infection.**

**Multicollinearity**

We produced generated the generalised variance inflation factor (GVIF) and adjusted generalised standard error inflation factor (aGSIF) to detect multicollinearity in models. No evidence of multicollinearity was detected, with all GVIF and aGSIF values below 10 and 2, respectively (see below Tables 3-5).

### **Supplementary Table 3. Generalised variance inflation factor (GVIF) and adjusted generalised standard error inflation factor (aGSIF) for regression models representing the total effect of IMD quintile on developing post-COVID condition by sex-at-birth and variant period in two analysis cohorts. GVIF of below 10 and aGSIF of below 2 suggest a lack of evidence of multicollinearity.**

**Exposure: IMD Quintile**

|  | **GVIF (aGSIF)** | | | | | | | |
| --- | --- | --- | --- | --- | --- | --- | --- | --- |
| **Analysis cohort** | Full cohort (regardless of previous SARS-CoV-2 infection) | | | | Individuals with a reported previous SARS-CoV-2 infection | | | |
| **Sex-at-birth** | Male | | Female | | Male | | Female | |
| **Time period** | Pre-Omicron | Omicron | Pre-Omicron | Omicron | Pre-Omicron | Omicron | Pre-Omicron | Omicron |
| IMD Quintile | 1.06 (1.01) | 1.06 (1.01) | 1.05 (1.00) | 1.05 (1.00) | 1.14 (1.01) | 1.08 (1.01) | 1.05 (1.01) | 1.06 (1.01) |
| Age group | 2.61 (1.27) | 2.47 (1.25) | 2.22 (1.22) | 2.34 (1.24) | 2.00 (1.19) | 1.80 (1.16) | 1.81 (1.16) | 1.65 (1.13) |
| Minority ethnicity status | 4.64 (1.21) | 4.31 (1.20) | 2.64 (1.13) | 2.29 (1.11) | 1.24 (1.03) | 1.19 (1.02) | 1.15 (1.02) | 1.15 (1.02) |
| Migration status | 1.85 (1.17) | 1.70 (1.14) | 1.53 (1.11) | 1.57 (1.12) | 2.03 (1.19) | 1.80 (1.16) | 1.74 (1.32) | 1.67 (1.14) |
| Occupation | 5.40 (1.23) | 5.29 (1.23) | 3.47 (1.17) | 3.18 (1.16) | - | - | - | - |

### **Supplementary Table 4. Generalised variance inflation factor (GVIF) and adjusted generalised standard error inflation factor (aGSIF) for regression models representing the total effect of migration status on developing post-COVID condition by sex-at-birth and variant period in two analysis cohorts. GVIF of below 10 and aGSIF of below 2 suggest a lack of evidence of multicollinearity.**

**Exposure: Migration status**

|  | **GVIF (aGSIF)** | | | | | | | |
| --- | --- | --- | --- | --- | --- | --- | --- | --- |
| **Analysis cohort** | Full cohort (regardless of previous SARS-CoV-2 infection) | | | | Individuals with a reported previous SARS-CoV-2 infection | | | |
| **Sex-at-birth** | Male | | Female | | Male | | Female | |
| **Time period** | Pre-Omicron | Omicron | Pre-Omicron | Omicron | Pre-Omicron | Omicron | Pre-Omicron | Omicron |
| Migration status | 1.75 (1.15) | 1.60 (1.13) | 1.48 (1.10) | 1.49 (1.11) | 1.76 (1.15) | 1.71 (1.14) | 1.78 (1.16) | 1.64 (1.13) |
| Age group | 1.12 (1.01) | 1.10 (1.01) | 1.10 (1.01) | 1.10 (1.01) | 1.20 (1.02) | 1.15 (1.02) | 1.12 (1.01) | 1.13 (1.02) |
| Minority ethnicity status | 1.76 (1.15) | 1.64 (1.13) | 1.50 (1.11) | 1.54 (1.11) | 1.87 (1.17) | 1.76 (1.15) | 1.73 (1.31) | 1.66 (1.14) |

### **Supplementary Table 5. Generalised variance inflation factor (GVIF) and adjusted generalised standard error inflation factor (aGSIF) for regression models representing the total effect of ethnic minority status on developing post-COVID condition by sex-at-birth and variant period in two analysis cohorts. GVIF of below 10 and an aGSIF of below 2 suggest a lack of evidence of multicollinearity.**

**Exposure: Minority ethnicity status**

|  | **GVIF (aGSIF)** | | | | | | | |
| --- | --- | --- | --- | --- | --- | --- | --- | --- |
| **Analysis cohort** | Full cohort (regardless of previous SARS-CoV-2 infection) | | | | Individuals with a reported previous SARS-CoV-2 infection | | | |
| **Sex-at-birth** | Male | | Female | | Male | | Female | |
| **Time period** | Pre-Omicron | Omicron | Pre-Omicron | Omicron | Pre-Omicron | Omicron | Pre-Omicron | Omicron |
| Minority ethnicity status | 1.06 (1.01) | 1.06 (1.01) | 1.06 (1.01) | 1.06 (1.01) | 1.13 (1.02) | 1.09 (1.01) | 1.05 (1.01) | 1.09 (1.01) |
| Age group | 1.06 (1.02) | 1.06 (1.01) | 1.6 (1.01) | 1.06 (1.02) | 1.13 (1.03) | 1.09 (1.02) | 1.05 (1.03) | 1.09 (1.02) |

**Supplementary Table 6. Sociodemographic and clinical characteristics of the analysis cohorts by IMD quintile.**

|  | **All participants regardless of any previous SARS-CoV-2 infection** | | | | | **Participants with a reported previous SARS-CoV-2 infection** | | | | |
| --- | --- | --- | --- | --- | --- | --- | --- | --- | --- | --- |
| **Characteristic** | **1**  **N = 593** | **2**  **N = 1,139** | **3**  **N = 1,734** | **4**  **N = 2,274** | **5**  **N = 2,778** | **1**  **N = 213** | **2**  **N = 406** | **3**  **N = 573** | **4**  **N = 714** | **5**  **N = 921** |
| **PCC** | 46 (7.8%) | 82 (7.2%) | 107 (6.2%) | 99 (4.4%) | 117 (4.2%) | 46 (22%) | 82 (20%) | 107 (19%) | 99 (14%) | 117 (13%) |
| **Age group** | | | | | | | | | | |
| 0-44* | 86 (15%) | 177 (16%) | 165 (9.5%) | 195 (8.6%) | 264 (9.5%) | 40 (19%) | 101 (25%) | 86 (15%) | 112 (16%) | 158 (17%) |
| 45-64 | 254 (43%) | 445 (39%) | 652 (38%) | 850 (37%) | 958 (34%) | 93 (44%) | 166 (41%) | 251 (44%) | 285 (40%) | 342 (37%) |
| 65+ | 253 (43%) | 517 (45%) | 917 (53%) | 1,229 (54%) | 1,556 (56%) | 80 (38%) | 139 (34%) | 236 (41%) | 317 (44%) | 421 (46%) |
| **Sex (at birth)** | | | | | | | | | | |
| Female | 345 (58%) | 673 (59%) | 998 (58%) | 1,286 (57%) | 1,562 (56%) | 125 (59%) | 258 (64%) | 331 (58%) | 404 (57%) | 528 (57%) |
| Male | 248 (42%) | 466 (41%) | 736 (42%) | 988 (43%) | 1,216 (44%) | 88 (41%) | 148 (36%) | 242 (42%) | 310 (43%) | 393 (43%) |
| **Ethnic minority status** | | | | | | | | | | |
| White British | 514 (87%) | 1,010 (89%) | 1,578 (91%) | 2,114 (93%) | 2,606 (94%) | 184 (86%) | 344 (85%) | 518 (90%) | 663 (93%) | 858 (93%) |
| Minority ethnic | 73 (12%) | 123 (11%) | 151 (8.7%) | 151 (6.6%) | 160 (5.8%) | 28 (13%) | 59 (15%) | 54 (9.4%) | 48 (6.7%) | 60 (6.5%) |
| Missing/Prefer not to say | 6 (1.0%) | 6 (0.5%) | 5 (0.3%) | 9 (0.4%) | 12 (0.4%) | 1 (0.5%) | 3 (0.7%) | 1 (0.2%) | 3 (0.4%) | 3 (0.3%) |
| **Migration status** | | | | | | | | | | |
| UK Born | 497 (84%) | 943 (83%) | 1,475 (85%) | 1,975 (87%) | 2,369 (85%) | 173 (81%) | 318 (78%) | 475 (83%) | 614 (86%) | 765 (83%) |
| Not UK Born | 48 (8.1%) | 99 (8.7%) | 118 (6.8%) | 126 (5.5%) | 140 (5.0%) | 21 (9.9%) | 40 (9.9%) | 33 (5.8%) | 32 (4.5%) | 44 (4.8%) |
| Missing | 48 (8.1%) | 97 (8.5%) | 141 (8.1%) | 173 (7.6%) | 269 (9.7%) | 19 (8.9%) | 48 (12%) | 65 (11%) | 68 (9.5%) | 112 (12%) |
| **Clinical vulnerability** | | | | | | | | | | |
| Not clinically vulnerable | 293 (49%) | 659 (58%) | 947 (55%) | 1,258 (55%) | 1,656 (60%) | 105 (49%) | 245 (60%) | 321 (56%) | 414 (58%) | 588 (64%) |
| Clinically vulnerable | 174 (29%) | 289 (25%) | 499 (29%) | 667 (29%) | 748 (27%) | 66 (31%) | 103 (25%) | 154 (27%) | 203 (28%) | 232 (25%) |
| Clinically extremely vulnerable | 121 (20%) | 181 (16%) | 274 (16%) | 313 (14%) | 363 (13%) | 41 (19%) | 52 (13%) | 90 (16%) | 87 (12%) | 97 (11%) |
| Missing | 5 (0.8%) | 10 (0.9%) | 14 (0.8%) | 36 (1.6%) | 11 (0.4%) | 1 (0.5%) | 6 (1.5%) | 8 (1.4%) | 10 (1.4%) | 4 (0.4%) |
| **Body Mass Index (BMI)** | | | | | | | | | | |
| Underweight/Normal | 161 (27%) | 360 (32%) | 591 (34%) | 776 (34%) | 1,014 (37%) | 62 (29%) | 125 (31%) | 176 (31%) | 230 (32%) | 324 (35%) |
| Pre-obesity | 154 (26%) | 328 (29%) | 487 (28%) | 711 (31%) | 840 (30%) | 59 (28%) | 103 (25%) | 158 (28%) | 218 (31%) | 267 (29%) |
| Obesity/Severe obesity | 149 (25%) | 223 (20%) | 343 (20%) | 404 (18%) | 407 (15%) | 53 (25%) | 78 (19%) | 112 (20%) | 128 (18%) | 133 (14%) |
| Missing | 129 (22%) | 228 (20%) | 313 (18%) | 383 (17%) | 517 (19%) | 39 (18%) | 100 (25%) | 127 (22%) | 138 (19%) | 197 (21%) |
| **Occupation** | | | | | | | | | | |
| Higher Risk Occupation | 117 (20%) | 219 (19%) | 304 (18%) | 352 (15%) | 385 (14%) | 55 (26%) | 101 (25%) | 131 (23%) | 143 (20%) | 160 (17%) |
| Lower Risk Occupation | 143 (24%) | 271 (24%) | 337 (19%) | 455 (20%) | 529 (19%) | 55 (26%) | 107 (26%) | 128 (22%) | 159 (22%) | 193 (21%) |
| Not in employment | 49 (8.3%) | 64 (5.6%) | 77 (4.4%) | 73 (3.2%) | 92 (3.3%) | 11 (5.2%) | 21 (5.2%) | 26 (4.5%) | 27 (3.8%) | 31 (3.4%) |
| Retired | 217 (37%) | 462 (41%) | 834 (48%) | 1,150 (51%) | 1,434 (52%) | 72 (34%) | 123 (30%) | 206 (36%) | 293 (41%) | 405 (44%) |
| Unknown or Other Status | 67 (11%) | 123 (11%) | 182 (10%) | 244 (11%) | 338 (12%) | 20 (9.4%) | 54 (13%) | 82 (14%) | 92 (13%) | 132 (14%) |
| n (%) | | | | | | | | | | |

*Age group collapsed to 0-44 to reduce the risk of disclosure.

**Supplementary Table 7. Sociodemographic and clinical characteristics of the analysis cohorts by migration status.**

|  | **All participants regardless of any previous SARS-CoV-2 infection** | | | **Participants with a reported previous SARS-CoV-2 infection** | | |
| --- | --- | --- | --- | --- | --- | --- |
| **Characteristic** | **UK Born,**  **N = 7,299** | **Not UK Born**  **N = 537** | **Missing**  **N = 755** | **UK Born**  **N = 2,355** | **Not UK Born**  **N = 171** | **Missing**  **N = 315** |
| **PCC** | 297 (4.1%) | 40 (7.4%) | 114 (15%) | 297 (13%) | 40 (23%) | 114 (36%) |
| **Age group** | | | | | | |
| 0-44* | 659 (9.0%) | 90 (17%) | 143 (19%) | 356 (15%) | 56 (33%) | 86 (27%) |
| 45-64 | 2,689 (37%) | 233 (43%) | 263 (35%) | 944 (40%) | 74 (43%) | 123 (39%) |
| 65+ | 3,951 (54%) | 214 (40%) | 349 (46%) | 1,055 (45%) | 41 (24%) | 106 (34%) |
| **Sex (at birth)** | | | | | | |
| Female | 4,191 (57%) | 324 (60%) | 391 (52%) | 1,369 (58%) | 100 (58%) | 186 (59%) |
| Male | 3,108 (43%) | 213 (40%) | 364 (48%) | 986 (42%) | 71 (42%) | 129 (41%) |
| **Ethnic minority status** | | | | | | |
| White British | 7,056 (97%) | 181 (34%) | 633 (84%) | 2,266 (96%) | 46 (27%) | 266 (84%) |
| Minority ethnic | 234 (3.2%) | 348 (65%) | 84 (11%) | 87 (3.7%) | 124 (73%) | 39 (12%) |
| Missing/Prefer not to say | 9 (0.1%) | 8 (1.5%) | 38 (5.0%) | 2 (<0.1%) | 1 (0.6%) | 10 (3.2%) |
| **IMD Quintile** | | | | | | |
| 1 | 497 (6.8%) | 48 (8.9%) | 48 (6.4%) | 173 (7.3%) | 21 (12%) | 19 (6.0%) |
| 2 | 943 (13%) | 99 (18%) | 97 (13%) | 318 (14%) | 40 (23%) | 48 (15%) |
| 3 | 1,475 (20%) | 118 (22%) | 141 (19%) | 475 (20%) | 33 (19%) | 65 (21%) |
| 4 | 1,975 (27%) | 126 (23%) | 173 (23%) | 614 (26%) | 32 (19%) | 68 (22%) |
| 5 | 2,369 (32%) | 140 (26%) | 269 (36%) | 765 (32%) | 44 (26%) | 112 (36%) |
| Missing | 40 (0.5%) | 6 (1.1%) | 27 (3.6%) | 10 (0.4%) | 1 (0.6%) | 3 (1.0%) |
| **Clinical vulnerability** | | | | | | |
| Not clinically vulnerable | 4,011 (55%) | 322 (60%) | 523 (69%) | 1,352 (57%) | 106 (62%) | 221 (70%) |
| Clinically vulnerable | 2,215 (30%) | 122 (23%) | 54 (7.2%) | 692 (29%) | 30 (18%) | 41 (13%) |
| Clinically extremely vulnerable | 1,067 (15%) | 92 (17%) | 103 (14%) | 310 (13%) | 34 (20%) | 26 (8.3%) |
| Missing | 6 (<0.1%) | 1 (0.2%) | 75 (9.9%) | 1 (<0.1%) | 1 (0.6%) | 27 (8.6%) |
| **Body Mass Index (BMI)** | | | | | | |
| Underweight/Normal | 2,666 (37%) | 221 (41%) | 31 (4.1%) | 816 (35%) | 75 (44%) | 31 (9.8%) |
| Pre-obesity | 2,347 (32%) | 170 (32%) | 18 (2.4%) | 748 (32%) | 42 (25%) | 18 (5.7%) |
| Obesity/Severe obesity | 1,428 (20%) | 89 (17%) | 18 (2.4%) | 457 (19%) | 32 (19%) | 18 (5.7%) |
| Missing | 858 (12%) | 57 (11%) | 688 (91%) | 334 (14%) | 22 (13%) | 248 (79%) |
| **Occupation** | | | | | | |
| Higher Risk Occupation | 1,190 (16%) | 124 (23%) | 72 (9.5%) | 496 (21%) | 49 (29%) | 46 (15%) |
| Lower Risk Occupation | 1,475 (20%) | 170 (32%) | 101 (13%) | 533 (23%) | 66 (39%) | 44 (14%) |
| Not in employment | 320 (4.4%) | 28 (5.2%) | 10 (1.3%) | 95 (4.0%) | 13 (7.6%) | 10 (3.2%) |
| Retired | 3,897 (53%) | 197 (37%) | 26 (3.4%) | 1,043 (44%) | 36 (21%) | 26 (8.3%) |
| Unknown or Other Status | 417 (5.7%) | 18 (3.4%) | 546 (72%) | 188 (8.0%) | 7 (4.1%) | 189 (60%) |
| n (%) | | | | | | |

*Age group collapsed to 0-44 to reduce the risk of disclosure.

**Supplementary Table 8. Sociodemographic and clinical characteristics of the analysis cohorts by ethnic minority status.**

|  | **All participants regardless of any previous SARS-CoV-2 infection** | | **Participants with a reported previous SARS-CoV-2 infection** | |
| --- | --- | --- | --- | --- |
| **Characteristic** | **White British**  **N = 7,870** | **Minority ethnic**  **N = 666** | **White British**  **N = 2,578** | **Minority ethnic**  **N = 250** |
| **PCC** | 391 (5.0%) | 56 (8.4%) | 391 (15%) | 56 (22%) |
| **Age group** | | | | |
| 0-44* | 686 (8.7%) | 201 (30%) | 376 (15%) | 119 (48%) |
| 45-64 | 2,878 (37%) | 281 (42%) | 1,051 (41%) | 84 (34%) |
| 65+ | 4,306 (55%) | 184 (28%) | 1,151 (45%) | 47 (19%) |
| **Sex (at birth)** | | | | |
| Female | 4,482 (57%) | 393 (59%) | 1,501 (58%) | 147 (59%) |
| Male | 3,388 (43%) | 273 (41%) | 1,077 (42%) | 103 (41%) |
| **Ethnic minority status** | | | | |
| White British | 7,056 (90%) | 234 (35%) | 2,266 (88%) | 87 (35%) |
| Minority ethnic | 181 (2.3%) | 348 (52%) | 46 (1.8%) | 124 (50%) |
| Missing/Prefer not to say | 633 (8.0%) | 84 (13%) | 266 (10%) | 39 (16%) |
| **IMD Quintile** | | | | |
| 1 | 514 (6.5%) | 73 (11%) | 184 (7.1%) | 28 (11%) |
| 2 | 1,010 (13%) | 123 (18%) | 344 (13%) | 59 (24%) |
| 3 | 1,578 (20%) | 151 (23%) | 518 (20%) | 54 (22%) |
| 4 | 2,114 (27%) | 151 (23%) | 663 (26%) | 48 (19%) |
| 5 | 2,606 (33%) | 160 (24%) | 858 (33%) | 60 (24%) |
| Missing | 48 (0.6%) | 8 (1.2%) | 11 (0.4%) | 1 (0.4%) |
| **Clinical vulnerability** | | | | |
| Not clinically vulnerable | 4,405 (56%) | 415 (62%) | 1,496 (58%) | 173 (69%) |
| Clinically vulnerable | 2,252 (29%) | 137 (21%) | 724 (28%) | 39 (16%) |
| Clinically extremely vulnerable | 1,145 (15%) | 110 (17%) | 334 (13%) | 36 (14%) |
| Missing | 68 (0.9%) | 4 (0.6%) | 24 (0.9%) | 2 (0.8%) |
| **Body Mass Index (BMI)** | | | | |
| Underweight/Normal | 2,682 (34%) | 231 (35%) | 834 (32%) | 86 (34%) |
| Pre-obesity | 2,374 (30%) | 156 (23%) | 762 (30%) | 46 (18%) |
| Obesity/Severe obesity | 1,435 (18%) | 99 (15%) | 472 (18%) | 35 (14%) |
| Missing | 1,379 (18%) | 180 (27%) | 510 (20%) | 83 (33%) |
| **Occupation** | | | | |
| Higher Risk Occupation | 1,243 (16%) | 135 (20%) | 530 (21%) | 59 (24%) |
| Lower Risk Occupation | 1,528 (19%) | 209 (31%) | 557 (22%) | 84 (34%) |
| Not in employment | 320 (4.1%) | 37 (5.6%) | 103 (4.0%) | 15 (6.0%) |
| Retired | 3,955 (50%) | 164 (25%) | 1,073 (42%) | 32 (13%) |
| Unknown or Other Status | 824 (10%) | 121 (18%) | 315 (12%) | 60 (24%) |
| n (%) | | | | |

*Age group collapsed to 0-44 to reduce the risk of disclosure.

**Supplementary Table 9. Unadjusted predicted probability and 95% confidence interval of developing PCC by IMD quintile, migration status and ethnic minority status in males and females during the pre-Omicron period by infection status.**

| **Pre-Omicron** | **Regardless of any previous SARS-CoV-2 infections** | | **With a history of SARS-CoV-2 infection** | |
| --- | --- | --- | --- | --- |
| **Sex-at-birth** | **Male** | **Female** | **Male** | **Female** |
| **IMD Quintile** | **(n = 2,859)** | **(n = 3,846)** | **(n = 400)** | **(n = 643)** |
| 1 (most deprived) | 4.12 (1.33, 6.92) | 7.78 (4.58, 10.97) | 22.22 (8.64, 35.80) | 38.89 (25.89, 51.89) |
| 2 | 4.24 (2.21, 6.28) | 6.82 (4.64, 9.00) | 26.67 (15.48, 37.86) | 35.00 (25.65, 44.35) |
| 3 | 3.46 (1.97, 4.95) | 5.47 (3.88, 7.06) | 23.81 (14.7, 32.92) | 34.96 (26.53, 43.39) |
| 4 | 1.56 (0.68, 2.43) | 5.16 (3.82, 6.50) | 12.37 (5.82, 18.92) | 32.53 (25.4, 39.66) |
| 5 (least deprived) | 1.81 (0.96, 2.66) | 3.50 (2.47, 4.52) | 13.82 (7.72, 19.92) | 21.50 (15.81, 27.19) |
| **Migration status** | **(n = 2,883)** | **(n = 3,878)** | **(n = 404)** | **(n = 651)** |
| UK-born | 1.87 (1.34, 2.41) | 4.1 (3.43, 4.78) | 13.65 (9.98, 17.32) | 26.25 (22.47, 30.02) |
| Non-UK-born | 7.19 (3.27, 11.1) | 6.06 (3.18, 8.94) | 44.44 (25.7, 63.19) | 35.56 (21.57, 49.54) |
| Missing | 5.75 (2.92, 8.57) | 15.64 (11.34, 19.93) | 37.5 (22.5, 52.5) | 51.19 (40.5, 61.88) |
| **Ethnic minority status** | **(n = 2,883)** | **(n = 3,878)** | **(n = 404)** | **(n = 651)** |
| White British | 2.24 (1.68, 2.80) | 4.74 (4.04, 5.44) | 16.04 (12.32, 19.76) | 28.84 (25.17, 32.51) |
| Ethnic minority | 6.50 (3.08, 9.92) | 8.68 (5.55, 11.81) | 43.33 (25.6, 61.07) | 41.54 (29.56, 53.52) |

**Supplementary Table 10. Unadjusted predicted probability and 95% confidence interval of developing PCC by IMD quintile, migration status and ethnic minority status in males and females during the Omicron period by infection status.**

| **Omicron** | **Regardless of any previous SARS-CoV-2 infections** | | **With a history of SARS-CoV-2 infection** | |
| --- | --- | --- | --- | --- |
| **Sex-at-birth** | **Male** | **Female** | **Male** | **Female** |
| **IMD Quintile** | **(n = 3,252)** | **(n = 4,234)** | **(n = 793)** | **(n = 1,031)** |
| 1 (most deprived) | 1.9 (0.06, 3.74) | 4.53 (2.12, 6.94) | 7.55 (0.44, 14.66) | 18.31 (9.31, 27.31) |
| 2 | 1.99 (0.62, 3.35) | 3.82 (2.25, 5.38) | 9.3 (3.16, 15.44) | 13.5 (8.25, 18.74) |
| 3 | 2.28 (1.14, 3.43) | 3.1 (1.95, 4.25) | 9.2 (4.76, 13.64) | 12.98 (8.41, 17.55) |
| 4 | 1.35 (0.59, 2.12) | 1.77 (1.00, 2.54) | 5.63 (2.54, 8.73) | 8 (4.64, 11.36) |
| 5 (least deprived) | 1.83 (1.03, 2.62) | 2.63 (1.78, 3.48) | 7.19 (4.16, 10.23) | 10.62 (7.34, 13.9) |
| **Migration status** | **(n = 3,276)** | **(n = 4,266)** | **(n = 797)** | **(n = 1,039)** |
| UK-born | 1.44 (0.99, 1.88) | 2.00 (1.55, 2.45) | 6 (4.2, 7.8) | 8.43 (6.59, 10.27) |
| Non-UK-born | 1.08 (0.00, 2.57) | 3.60 (1.41, 5.79) | 4.44 (0.00, 10.47) | 16.95 (7.38, 26.52) |
| Missing | 5.56 (2.99, 8.12) | 11.6 (7.94, 15.27) | 20 (11.5, 28.5) | 33.33 (24.19, 42.48) |
| **Ethnic minority status** | **(n = 3,276)** | **(n = 4,266)** | **(n = 797)** | **(n = 1,039)** |
| White British | 1.85 (1.37, 2.33) | 2.67 (2.17, 3.17) | 7.75 (5.8, 9.69) | 11.03 (9.04, 13.02) |
| Ethnic minority | 1.23 (0.00, 2.61) | 3.9 (1.82, 5.98) | 4.05 (0.00, 8.55) | 14.94 (7.45, 22.43) |

**Supplementary Table 11. Adjusted predicted probability and 95% confidence interval of developing PCC by IMD quintile, migration status and ethnic minority status in males and females during the Pre-Omicron period by infection status.**

| **Pre-Omicron** | **Regardless of any previous SARS-CoV-2 infections** | | **With a history of SARS-CoV-2 infection** | |
| --- | --- | --- | --- | --- |
| **Sex-at-birth** | **Male** | **Female** | **Male** | **Female** |
| **IMD Quintile** | **(n = 2,859)** | **(n = 3,846)** | **(n = 400)** | **(n = 643)** |
| 1 (most deprived) | 3.68 (1.23, 6.14) | 6.22 (3.73, 8.7) | 19.15 (7.31, 30.99) | 37.74 (25.40, 50.07) |
| 2 | 3.5 (1.84, 5.16) | 6.18 (4.26, 8.11) | 26.1 (15.54, 36.66) | 32.65 (23.87, 41.43) |
| 3 | 3.42 (1.97, 4.87) | 5.49 (3.95, 7.02) | 24.31 (15.57, 33.06) | 35.5 (27.33, 43.67) |
| 4 | 1.7 (0.76, 2.64) | 5.34 (4.00, 6.67) | 13.77 (6.98, 20.55) | 32.12 (25.32, 38.92) |
| 5 (least deprived) | 1.92 (1.03, 2.8) | 3.77 (2.70, 4.84) | 13.33 (7.56, 19.09) | 22.58 (16.83, 28.32) |
| **Migration status** | **(n = 2,883)** | **(n = 3,878)** | **(n = 404)** | **(n = 651)** |
| UK-born | 1.92 (1.37, 2.48) | 4.24 (3.54, 4.93) | 14.42 (10.50, 18.34) | 27.06 (23.19, 30.94) |
| Non-UK-born | 6.33 (1.86, 10.8) | 4.76 (2.09, 7.42) | 33.14 (11.79, 54.50) | 23.78 (10.06, 37.5) |
| Missing | 5.00 (2.54, 7.47) | 14.26 (10.34, 18.18) | 34.22 (20.05, 48.40) | 53.76 (43.21, 64.32) |
| **Ethnic minority status** | **(n = 2,883)** | **(n = 3,878)** | **(n = 404)** | **(n = 651)** |
| White British | 2.33 (1.75, 2.90) | 4.97 (4.25, 5.69) | 16.24 (12.51, 19.96) | 29.19 (25.55, 32.84) |
| Ethnic minority | 4.33 (1.97, 6.69) | 5.65 (3.52, 7.78) | 38.8 (21.17, 56.43) | 38.01 (26.24, 49.79) |

**Supplementary Table 12. Adjusted predicted probability and 95% confidence interval of developing PCC by IMD quintile, migration status and ethnic minority status in males and females during the Omicron period by infection status.**

| **Omicron** | **Regardless of any previous SARS-CoV-2 infections** | | **With a history of SARS-CoV-2 infection** | |
| --- | --- | --- | --- | --- |
| **Sex-at-birth** | **Male** | **Female** | **Male** | **Female** |
| **IMD Quintile** | **(n =3,252)** | **(n = 4,234)** | **(n = 793)** | **(n = 1031)** |
| 1 (most deprived) | 2.14 (0.09, 4.19) | 4.20 (2.04, 6.36) | 8.20 (0.65, 15.76) | 18.08 (9.58, 26.59) |
| 2 | 1.95 (0.62, 3.28) | 3.46 (2.06, 4.87) | 9.62 (3.40, 15.85) | 12.39 (7.57, 17.2) |
| 3 | 2.4 (1.21, 3.59) | 3.11 (2.00, 4.23) | 9.02 (4.70, 13.35) | 12.93 (8.57, 17.29) |
| 4 | 1.37 (0.61, 2.13) | 1.83 (1.05, 2.60) | 6.04 (2.78, 9.30) | 8.27 (4.91, 11.62) |
| 5 (least deprived) | 1.73 (0.98, 2.48) | 2.72 (1.87, 3.57) | 6.76 (3.91, 9.60) | 10.89 (7.65, 14.14) |
| **Migration status** | **(n = 3,276)** | **(n = 4,266)** | **(n = 797)** | **(n = 1039)** |
| UK-born | 1.41 (0.98, 1.85) | 2.00 (1.55, 2.46) | 5.83 (4.07, 7.59) | 8.41 (6.56, 10.25) |
| Non-UK-born | 1.56 (0.00, 3.93) | 3.57 (1.03, 6.11) | 8.01 (0.00, 19.77) | 16.09 (4.96, 27.21) |
| Missing | 5.43 (2.89, 7.96) | 11.83 (8.09, 15.58) | 19.79 (11.22, 28.36) | 35.14 (25.75, 44.53) |
| **Ethnic minority status** | **(n = 3,276)** | **(n = 4,266)** | **(n = 797)** | **(n = 1039)** |
| White British | 1.87 (1.38, 2.35) | 2.73 (2.21, 3.24) | 7.78 (5.82, 9.74) | 10.98 (9.00, 12.96) |
| Ethnic minority | 1.08 (0.00, 2.31) | 3.15 (1.41, 4.90) | 3.88 (0.00, 8.26) | 15.68 (7.53, 23.84) |

**Supplementary Table 13. Unadjusted odds ratio (OR) and adjusted odds ratio (aOR) for the association between IMD quintile and developing PCC in males during the Pre-Omicron period by infection status.**

| **Pre-Omicron**  **Sex = Male** | **Regardless of any previous SARS-CoV-2 infections**  **(n = 2,859)** | | | | **With a history of SARS-CoV-2 infection**  **(n = 400)** | | | |
| --- | --- | --- | --- | --- | --- | --- | --- | --- |
| **Variable** | **OR** | **95 CI** | **aOR** | **95 CI** | **OR** | **95 CI** | **aOR** | **95 CI** |
| **IMD Quintile** | | | | | | | | |
| 1 (most deprived) | 2.34 | 0.99, 5.49 | 2.02 | 0.83, 4.88 | 1.78 | 0.70, 4.55 | 1.61 | 0.59, 4.44 |
| 2 | 2.41 | 1.20, 4.81 | 1.91 | 0.93, 3.89 | 2.27 | 1.05, 4.89 | 2.53 | 1.09, 5.84 |
| 3 | 1.95 | 1.01, 3.75 | 1.86 | 0.95, 3.65 | 1.95 | 0.95, 3.99 | 2.27 | 1.04, 4.93 |
| 4 | 0.86 | 0.41, 1.81 | 0.88 | 0.41, 1.88 | 0.88 | 0.40, 1.94 | 1.04 | 0.45, 2.40 |
| 5 (least deprived) | ref | - | ref | - | ref | - | ref | - |
| **Age group** | | | | | | | | |
| 0-24 | - | - | 2.45 | 1.06, 5.65 | - | - | 0.79 | 0.36, 1.78 |
| 25-44 | - | - | 1.56 | 0.74, 3.28 | - | - | 1.00 | 0.40, 2.49 |
| 45-64 | - | - | ref | - | - | - | ref | - |
| 65+ | - | - | 0.25 | 0.12, 0.54 | - | - | 0.40 | 0.20, 0.82 |
| **Migration status** | | | | | | | | |
| UK Born | - | - | ref | - | - | - | ref | - |
| Non UK born | - | - | 3.05 | 1.24, 7.53 | - | - | 4.28 | 1.31, 14.00 |
| Missing | - | - | 4.49 | 2.09, 9.64 | - | - | 3.70 | 1.71, 7.99 |
| **Minority ethnicity status** | | | | | | | | |
| White British | - | - | ref | - | - | - | ref | - |
| Ethnic minority | - | - | 0.99 | 0.42, 2.34 | - | - | 1.36 | 0.42, 4.41 |
| **Occupation** | | | | | | | | |
| Higher risk occupation | - | - | ref | - | - | - | - | - |
| Lower risk occupation | - | - | 0.78 | 0.41, 1.46 | - | - | - | - |
| Not in employment | - | - | 0.47 | 0.20, 1.09 | - | - | - | - |
| Retired | - | - | 0.76 | 0.26, 2.22 | - | - | - | - |
| Unknown/Other status | - | - | 0.29 | 0.11, 0.77 | - | - | - | - |

**Supplementary Table 14. Unadjusted odds ratio (OR) and adjusted odds ratio (aOR) for the association between IMD quintile and developing PCC in females during the Pre-Omicron period by infection status.**

| **Pre-Omicron**  **Sex = Female** | **Regardless of any previous SARS-CoV-2 infections (n = 3,846)** | | | | **With a history of SARS-CoV-2 infection**  **(n = 643)** | | | |
| --- | --- | --- | --- | --- | --- | --- | --- | --- |
| **Variable** | **OR** | **95 CI** | **aOR** | **95 CI** | **OR** | **95 CI** | **aOR** | **95 CI** |
| **IMD Quintile** | | | | | | | | |
| 1 (most deprived) | 2.33 | 1.36, 3.99 | 1.76 | 1.00, 3.10 | 2.32 | 1.22, 4.42 | 2.20 | 1.13, 4.30 |
| 2 | 2.02 | 1.28, 3.20 | 1.75 | 1.08, 2.84 | 1.97 | 1.16, 3.35 | 1.73 | 0.99, 3.02 |
| 3 | 1.60 | 1.04, 2.46 | 1.53 | 0.97, 2.39 | 1.96 | 1.19, 3.24 | 1.98 | 1.18, 3.35 |
| 4 | 1.50 | 1.00, 2.26 | 1.48 | 0.97, 2.26 | 1.76 | 1.10, 2.81 | 1.68 | 1.03, 2.74 |
| 5 (least deprived) | ref | - | ref | - | ref | - | ref | - |
| **Age group** | | | | | | | | |
| 0-24 | - | - | 1.64 | 0.82, 3.29 | - | - | 0.34 | 0.17, 0.67 |
| 25-44 | - | - | 2.67 | 1.72, 4.13 | - | - | 1.50 | 0.89, 2.52 |
| 45-64 | - | - | ref | - | - | - | ref | - |
| 65+ | - | - | 0.39 | 0.25, 0.61 | - | - | 0.53 | 0.34, 0.82 |
| **Migration status** | | | | | | | | |
| UK Born | - | - | ref | - | - | - | ref | - |
| Non UK born | - | - | 1.09 | 0.56, 2.10 | - | - | 0.84 | 0.36, 1.98 |
| Missing | - | - | 7.80 | 4.75, 12.82 | - | - | 3.45 | 2.08, 5.72 |
| **Minority ethnicity status** | | | | | | | | |
| White British | - | - | ref | - | - | - | ref | - |
| Ethnic minority | - | - | 1.00 | 0.57, 1.74 | - | - | 1.51 | 0.73, 3.12 |
| **Occupation** | | | | | | | | |
| Higher risk occupation | - | - | ref | - | - | - | - | - |
| Lower risk occupation | - | - | 0.55 | 0.37, 0.81 | - | - | - | - |
| Not in employment | - | - | 0.46 | 0.28, 0.74 | - | - | - | - |
| Retired | - | - | 0.63 | 0.35, 1.15 | - | - | - | - |
| Unknown/Other status | - | - | 0.16 | 0.08, 0.32 | - | - | - | - |

**Supplementary Table 15. Unadjusted odds ratio (OR) and adjusted odds ratio (aOR) for the association between IMD quintile and developing PCC in males during the Omicron period by infection status.**

| **Omicron**  **Sex = Male** | **Regardless of any previous SARS-CoV-2 infections**  **(n = 3,252)** | | | | **With a history of SARS-CoV-2 infection**  **(n = 793)** | | | |
| --- | --- | --- | --- | --- | --- | --- | --- | --- |
| **Variable** | **OR** | **95 CI** | **aOR** | **95 CI** | **OR** | **95 CI** | **aOR** | **95 CI** |
| **IMD Quintile** | | | | | | | | |
| 1 (most deprived) | 1.04 | 0.35, 3.07 | 1.25 | 0.41, 3.77 | 1.05 | 0.34, 3.21 | 1.24 | 0.40, 3.90 |
| 2 | 1.09 | 0.48, 2.49 | 1.13 | 0.49, 2.64 | 1.32 | 0.56, 3.12 | 1.49 | 0.62, 3.60 |
| 3 | 1.26 | 0.64, 2.47 | 1.41 | 0.70, 2.81 | 1.31 | 0.65, 2.63 | 1.38 | 0.67, 2.85 |
| 4 | 0.74 | 0.36, 1.52 | 0.78 | 0.38, 1.63 | 0.77 | 0.37, 1.61 | 0.88 | 0.42, 1.88 |
| 5 (least deprived) | ref | - | ref | - | ref | - | ref | - |
| **Age group** | | | | | | | | |
| 0-24 | - | - | 2.83 | 0.98, 8.20 | - | - | 1.17 | 0.43, 3.18 |
| 25-44 | - | - | 0.70 | 0.16, 3.15 | - | - | 0.52 | 0.11, 2.40 |
| 45-64 | - | - | ref | - | - | - | ref | - |
| 65+ | - | - | 0.59 | 0.30, 1.13 | - | - | 0.75 | 0.41, 1.35 |
| **Migration status** | | | | | | | | |
| UK Born | - | - | ref | - | - | - | ref | - |
| Non UK born | - | - | 1.17 | 0.23, 5.90 | - | - | 1.61 | 0.30, 8.64 |
| Missing | - | - | 10.18 | 4.49, 23.07 | - | - | 3.97 | 2.09, 7.55 |
| **Minority ethnicity status** | | | | | | | | |
| White British | - | - | ref | - | - | - | ref | - |
| Ethnic minority | - | - | 0.50 | 0.13, 1.94 | - | - | 0.34 | 0.08, 1.38 |
| **Occupation** | | | | | | | | |
| Higher risk occupation | - | - | ref | - | - | - | - | - |
| Lower risk occupation | - | - | 0.62 | 0.26, 1.45 | - | - | - | - |
| Not in employment | - | - | 1.16 | 0.49, 2.76 | - | - | - | - |
| Retired | - | - | 0.40 | 0.05, 3.36 | - | - | - | - |
| Unknown/Other status | - | - | 0.20 | 0.07, 0.58 | - | - | - | - |

**Supplementary Table 16. Unadjusted odds ratio (OR) and adjusted odds ratio (aOR) for the association between IMD quintile and developing PCC in females during the Omicron period by infection status.**

| **Omicron**  **Sex = Female** | **Regardless of any previous SARS-CoV-2 infections (n = 4,234)** | | | | **With a history of SARS-CoV-2 infection**  **(n = 1,031)** | | | |
| --- | --- | --- | --- | --- | --- | --- | --- | --- |
| **Variable** | **OR** | **95 CI** | **aOR** | **95 CI** | **OR** | **95 CI** | **aOR** | **95 CI** |
| **IMD Quintile** | | | | | | | | |
| 1 (most deprived) | 1.76 | 0.92, 3.36 | 1.62 | 0.82, 3.18 | 1.89 | 0.94, 3.77 | 1.90 | 0.92, 3.94 |
| 2 | 1.47 | 0.86, 2.52 | 1.31 | 0.74, 2.30 | 1.31 | 0.75, 2.31 | 1.17 | 0.64, 2.14 |
| 3 | 1.18 | 0.71, 1.97 | 1.16 | 0.69, 1.96 | 1.26 | 0.74, 2.14 | 1.23 | 0.71, 2.15 |
| 4 | 0.67 | 0.38, 1.16 | 0.65 | 0.37, 1.15 | 0.73 | 0.41, 1.30 | 0.72 | 0.40, 1.30 |
| 5 (least deprived) | ref | - | ref | - | ref | - | ref | - |
| **Age group** | | | | | | | | |
| 0-24 | - | - | 0.19 | 0.03, 1.50 | - | - | 0.08 | 0.01, 0.62 |
| 25-44 | - | - | 1.56 | 0.80, 3.04 | - | - | 0.76 | 0.38, 1.51 |
| 45-64 | - | - | ref | - | - | - | ref | - |
| 65+ | - | - | 0.66 | 0.41, 1.08 | - | - | 0.66 | 0.43, 1.02 |
| **Migration status** | | | | | | | | |
| UK Born | - | - | ref | - | - | - | ref | - |
| Non UK born | - | - | 1.74 | 0.78, 3.88 | - | - | 1.96 | 0.80, 4.79 |
| Missing | - | - | 19.34 | 10.89, 34.35 | - | - | 6.26 | 3.80, 10.29 |
| **Minority ethnicity status** | | | | | | | | |
| White British | - | - | ref | - | - | - | ref | - |
| Ethnic minority | - | - | 0.84 | 0.39, 1.79 | - | - | 0.99 | 0.43, 2.26 |
| **Occupation** | | | | | | | | |
| Higher risk occupation | - | - | ref | - | - | - | - | - |
| Lower risk occupation | - | - | 0.62 | 0.35, 1.09 | - | - | - | - |
| Not in employment | - | - | 0.76 | 0.42, 1.36 | - | - | - | - |
| Retired | - | - | 0.96 | 0.42, 2.18 | - | - | - | - |
| Unknown/Other status | - | - | 0.12 | 0.05, 0.27 | - | - | - | - |

**Supplementary Table 17. Unadjusted odds ratio (OR) and adjusted odds ratio (aOR) for the association between migration status and developing PCC in males during the Pre-Omicron period by infection status.**

| **Pre-Omicron**  **Sex = Male** | **Regardless of any previous SARS-CoV-2 infections**  **(n = 2,883)** | | | | **With a history of SARS-CoV-2 infection**  **(n = 404)** | | | |
| --- | --- | --- | --- | --- | --- | --- | --- | --- |
| **Variable** | **OR** | **95 CI** | **aOR** | **95 CI** | **OR** | **95 CI** | **aOR** | **95 CI** |
| **Migration status** | | | | | | | | |
| UK born | ref | - | ref | - | ref | - | ref | - |
| Non UK born | 4.05 | 2.10, 7.81 | 3.57 | 1.49, 8.53 | 5.06 | 2.23, 11.49 | 3.07 | 1.03, 9.18 |
| Missing | 3.19 | 1.76, 5.80 | 2.75 | 1.48, 5.11 | 3.80 | 1.86, 7.73 | 3.23 | 1.54, 6.75 |
| **Age group** | | | | | | | | |
| 0-24 | - | - | 1.75 | 0.86, 3.55 | - | - | 0.79 | 0.36, 1.72 |
| 25-44 | - | - | 1.79 | 0.87, 3.69 | - | - | 1.02 | 0.41, 2.50 |
| 45-64 | - | - | ref | - | - | - | ref | - |
| 65+ | - | - | 0.18 | 0.09, 0.35 | - | - | 0.36 | 0.18, 0.73 |
| **Minority ethnicity status** | | | | | | | | |
| White British | - | - | ref | - | - | - | ref | - |
| Ethnic minority | - | - | 0.96 | 0.42, 2.21 | - | - | 1.70 | 0.56, 5.11 |

**Supplementary Table 18. Unadjusted odds ratio (OR) and adjusted odds ratio (aOR) for the association between migration status and developing PCC in females during the Pre-Omicron period by infection status.**

| **Pre-Omicron**  **Sex = Female** | **Regardless of any previous SARS-CoV-2 infections**  **(n = 3,878)** | | | | **With a history of SARS-CoV-2 infection**  **(n = 651)** | | | |
| --- | --- | --- | --- | --- | --- | --- | --- | --- |
| **Variable** | **OR** | **95 CI** | **aOR** | **95 CI** | **OR** | **95 CI** | **aOR** | **95 CI** |
| **Migration status** | | | | | | | | |
| UK born | ref | - | ref | - | ref | - | ref | - |
| Non UK born | 1.51 | 0.88, 2.57 | 1.13 | 0.59, 2.17 | 1.55 | 0.82, 2.94 | 0.83 | 0.36, 1.95 |
| Missing | 4.33 | 3.00, 6.26 | 4.02 | 2.72, 5.95 | 2.95 | 1.84, 4.72 | 3.34 | 2.03, 5.51 |
| **Age group** | | | | | | | | |
| 0-24 | - | - | 0.97 | 0.52, 1.81 | - | - | 0.32 | 0.16, 0.64 |
| 25-44 | - | - | 3.03 | 1.98, 4.63 | - | - | 1.50 | 0.90, 2.50 |
| 45-64 | - | - | ref | - | - | - | ref | - |
| 65+ | - | - | 0.27 | 0.19, 0.40 | - | - | 0.51 | 0.33, 0.79 |
| **Minority ethnicity status** | | | | | | | | |
| White British | - | - | ref | - | - | - | ref | - |
| Ethnic minority | - | - | 0.98 | 0.57, 1.68 | - | - | 1.65 | 0.80, 3.40 |

**Supplementary Table 19. Unadjusted odds ratio (OR) and adjusted odds ratio (aOR) for the association between migration status and developing PCC in males during the Omicron period by infection status.**

| **Omicron**  **Sex = Male** | **Regardless of any previous SARS-CoV-2 infections**  **(n = 3,276)** | | | | **With a history of SARS-CoV-2 infection**  **(n = 797)** | | | |
| --- | --- | --- | --- | --- | --- | --- | --- | --- |
| **Variable** | **OR** | **95 CI** | **aOR** | **95 CI** | **OR** | **95 CI** | **aOR** | **95 CI** |
| **Migration status** | | | | | | | | |
| UK born | ref | - | ref | - | ref | - | ref | - |
| Non UK born | 0.75 | 0.18, 3.13 | 1.11 | 0.23, 5.43 | 0.73 | 0.17, 3.12 | 1.41 | 0.27, 7.33 |
| Missing | 4.04 | 2.26, 7.21 | 4.02 | 2.23, 7.24 | 3.92 | 2.11, 7.29 | 4.03 | 2.13, 7.62 |
| **Age group** | | | | | | | | |
| 0-24 | - | - | 1.49 | 0.58, 3.79 | - | - | 1.10 | 0.41, 2.97 |
| 25-44 | - | - | 0.62 | 0.14, 2.69 | - | - | 0.54 | 0.12, 2.46 |
| 45-64 | - | - | ref | - | - | - | ref | - |
| 65+ | - | - | 0.64 | 0.36, 1.12 | - | - | 0.73 | 0.41, 1.32 |
| **Minority ethnicity status** | | | | | | | | |
| White British | - | - | ref | - | - | - | ref | - |
| Ethnic minority | - | - | 0.52 | 0.14, 1.94 | - | - | 0.37 | 0.09, 1.48 |

**Supplementary Table 20. Unadjusted odds ratio (OR) and adjusted odds ratio (aOR) for the association between migration status and developing PCC in females during the Omicron period by infection status.**

| **Omicron**  **Sex = Female** | **Regardless of any previous SARS-CoV-2 infections**  **(n = 4,266)** | | | | **With a history of SARS-CoV-2 infection**  **(n = 1,039)** | | | |
| --- | --- | --- | --- | --- | --- | --- | --- | --- |
| **Variable** | **OR** | **95 CI** | **aOR** | **95 CI** | **OR** | **95 CI** | **aOR** | **95 CI** |
| **Migration status** | | | | | | | | |
| UK born | ref | - | ref | - | ref | - | ref | - |
| Non UK born | 1.83 | 0.93, 3.57 | 1.82 | 0.82, 4.02 | 2.22 | 1.08, 4.56 | 2.10 | 0.86, 5.12 |
| Missing | 6.42 | 4.20, 9.83 | 6.68 | 4.32, 10.33 | 5.43 | 3.38, 8.74 | 6.08 | 3.72, 9.95 |
| **Age group** | | | | | | | | |
| 0-24 | - | - | 0.14 | 0.02, 1.06 | - | - | 0.08 | 0.01, 0.59 |
| 25-44 | - | - | 1.68 | 0.89, 3.15 | - | - | 0.78 | 0.39, 1.53 |
| 45-64 | - | - | ref | - | - | - | ref | - |
| 65+ | - | - | 0.54 | 0.36, 0.81 | - | - | 0.66 | 0.43, 1.01 |
| **Minority ethnicity status** | | | | | | | | |
| White British | - | - | ref | - | - | - | ref | - |
| Ethnic minority | - | - | 0.78 | 0.38, 1.62 | - | - | 1.05 | 0.46, 2.40 |

**Supplementary Table 21. Unadjusted odds ratio (OR) and adjusted odds ratio (aOR) for the association between ethnic minority status and developing PCC in males during the Pre-Omicron period by infection status.**

| **Pre-Omicron**  **Sex = Male** | **Regardless of any previous SARS-CoV-2 infections**  **(n = 2,883)** | | | | **With a history of SARS-CoV-2 infection**  **(n = 404)** | | | |
| --- | --- | --- | --- | --- | --- | --- | --- | --- |
| **Variable** | **OR** | **95 CI** | **aOR** | **95 CI** | **OR** | **95 CI** | **aOR** | **95 CI** |
| **Minority ethnicity status** | | | | | | | | |
| White British | ref | - | ref | - | ref | - | ref | - |
| Ethnic minority | 3.04 | 1.64, 5.64 | 1.93 | 1.01, 3.68 | 4.00 | 1.85, 8.67 | 3.44 | 1.49, 7.94 |
| **Age group** | | | | | | | | |
| 0-24 | - | - | 1.67 | 0.83, 3.34 | - | - | 0.77 | 0.36, 1.65 |
| 25-44 | - | - | 1.88 | 0.92, 3.84 | - | - | 1.02 | 0.43, 2.43 |
| 45-64 | - | - | ref | - | - | - | ref | - |
| 65+ | - | - | 0.18 | 0.09, 0.35 | - | - | 0.33 | 0.16, 0.66 |

**Supplementary Table 22. Unadjusted odds ratio (OR) and adjusted odds ratio (aOR) for the association between ethnic minority status and developing PCC in females during the Pre-Omicron period by infection status.**

| **Pre-Omicron**  **Sex = Female** | **Regardless of any previous SARS-CoV-2 infections**  **(n = 3,878)** | | | | **With a history of SARS-CoV-2 infection**  **(n = 651)** | | | |
| --- | --- | --- | --- | --- | --- | --- | --- | --- |
| **Variable** | **OR** | **95 CI** | **aOR** | **95 CI** | **OR** | **95 CI** | **aOR** | **95 CI** |
| **Minority ethnicity status** | | | | | | | | |
| White British | ref | - | ref | - | ref | - | ref | - |
| Ethnic minority | 1.91 | 1.25, 2.92 | 1.15 | 0.74, 1.79 | 1.75 | 1.04, 2.96 | 1.51 | 0.87, 2.62 |
| **Age group** | | | | | | | | |
| 0-24 | - | - | 1.29 | 0.70, 2.35 | - | - | 0.43 | 0.22, 0.82 |
| 25-44 | - | - | 3.05 | 2.01, 4.62 | - | - | 1.49 | 0.90, 2.47 |
| 45-64 | - | - | ref | - | - | - | ref | - |
| 65+ | - | - | 0.27 | 0.19, 0.40 | - | - | 0.53 | 0.35, 0.82 |

**Supplementary Table 23. Unadjusted odds ratio (OR) and adjusted odds ratio (aOR) for the association between ethnic minority status and developing PCC in males during the Omicron period by infection status.**

| **Omicron**  **Sex = Male** | **Regardless of any previous SARS-CoV-2 infections**  **(n = 3,276)** | | | | **With a history of SARS-CoV-2 infection**  **(n = 797)** | | | |
| --- | --- | --- | --- | --- | --- | --- | --- | --- |
| **Variable** | **OR** | **95 CI** | **aOR** | **95 CI** | **OR** | **95 CI** | **aOR** | **95 CI** |
| **Minority ethnicity status** | | | | | | | | |
| White British | ref | - | ref | - | ref | - | ref | - |
| Ethnic minority | 0.66 | 0.21, 2.13 | 0.57 | 0.18, 1.88 | 0.50 | 0.15, 1.65 | 0.48 | 0.14, 1.61 |
| **Age group** | | | | | | | | |
| 0-24 | - | - | 1.77 | 0.71, 4.44 | - | - | 1.24 | 0.47, 3.23 |
| 25-44 | - | - | 0.68 | 0.16, 2.93 | - | - | 0.50 | 0.11, 2.22 |
| 45-64 | - | - | ref | - | - | - | ref | - |
| 65+ | - | - | 0.63 | 0.36, 1.10 | - | - | 0.66 | 0.37, 1.17 |

**Supplementary Table 24. Unadjusted odds ratio (OR) and adjusted odds ratio (aOR) for the association between ethnic minority status and developing PCC in females during the Omicron period by infection status.**

| **Omicron**  **Sex = Female** | **Regardless of any previous SARS-CoV-2 infections**  **(n = 4,266)** | | | | **With a history of SARS-CoV-2 infection**  **(n = 1,039)** | | | |
| --- | --- | --- | --- | --- | --- | --- | --- | --- |
| **Variable** | **OR** | **95 CI** | **aOR** | **95 CI** | **OR** | **95 CI** | **aOR** | **95 CI** |
| **Minority ethnicity status** | | | | | | | | |
| White British | ref | - | ref | - | ref | - | ref | - |
| Ethnic minority | 1.48 | 0.82, 2.67 | 1.16 | 0.63, 2.14 | 1.42 | 0.76, 2.64 | 1.51 | 0.78, 2.93 |
| **Age group** | | | | | | | | |
| 0-24 | - | - | 0.20 | 0.03, 1.48 | - | - | 0.11 | 0.01, 0.80 |
| 25-44 | - | - | 1.82 | 0.99, 3.37 | - | - | 0.91 | 0.47, 1.73 |
| 45-64 | - | - | ref | - | - | - | ref | - |
| 65+ | - | - | 0.55 | 0.37, 0.82 | - | - | 0.68 | 0.45, 1.03 |

**Supplementary Table 25. Sensitivity Analyses - Unadjusted odds ratio (OR) and adjusted odds ratio (aOR) for the association between migration status and developing PCC in males during the Pre-Omicron period by infection status.**

| **Pre-Omicron**  **Sex = Male** | **Regardless of any previous SARS-CoV-2 infections**  **(n = 2,883)** | | | | **With a history of SARS-CoV-2 infection**  **(n = 404)** | | | |
| --- | --- | --- | --- | --- | --- | --- | --- | --- |
| **Variable** | **OR** | **95 CI** | **aOR** | **95 CI** | **OR** | **95 CI** | **aOR** | **95 CI** |
| **Migration status** | | | | | | | | |
| UK born | ref | - | ref | - | ref | - | ref | - |
| Non UK born | 3.39 | 1.79, 6.44 | 2.62 | 1.04, 6.64 | 4.16 | 1.85, 9.33 | 1.77 | 0.54, 5.81 |
| Missing | 1.83 | 0.24, 13.73 | 0.73 | 0.08, 6.65 | 1.30 | 0.14, 11.83 | 0.33 | 0.03, 4.05 |
| **Age group** | | | | | | | | |
| 0-24 | - | - | 1.90 | 0.94, 3.83 | - | - | 0.83 | 0.39, 1.78 |
| 25-44 | - | - | 1.96 | 0.95, 4.02 | - | - | 1.10 | 0.46, 2.66 |
| 45-64 | - | - | ref | - | - | - | ref | - |
| 65+ | - | - | 0.18 | 0.09, 0.35 | - | - | 0.33 | 0.17, 0.67 |
| **Minority ethnicity status** | | | | | | | | |
| White British | - | - | ref | - | - | - | ref | - |
| Ethnic minority | - | - | 1.17 | 0.46, 2.95 | - | - | 2.84 | 0.83, 9.72 |

**Supplementary Table 26. Sensitivity Analyses - Unadjusted odds ratio (OR) and adjusted odds ratio (aOR) for the association between migration status and developing PCC in females during the Pre-Omicron period by infection status.**

| **Pre-Omicron**  **Sex = Female** | **Regardless of any previous SARS-CoV-2 infections**  **(n = 3,878)** | | | | **With a history of SARS-CoV-2 infection**  **(n = 651)** | | | |
| --- | --- | --- | --- | --- | --- | --- | --- | --- |
| **Variable** | **OR** | **95 CI** | **aOR** | **95 CI** | **OR** | **95 CI** | **aOR** | **95 CI** |
| **Migration status** | | | | | | | | |
| UK born | ref | - | ref | - | ref | - | ref | - |
| Non UK born | 1.26 | 0.74, 2.14 | 0.88 | 0.45, 1.76 | 1.34 | 0.71, 2.53 | 0.76 | 0.31, 1.86 |
| Missing | 4.05 | 1.66, 9.88 | 1.83 | 0.62, 5.45 | 3.64 | 1.01, 13.05 | 2.33 | 0.51, 10.76 |
| **Age group** | | | | | | | | |
| 0-24 | - | - | 1.25 | 0.68, 2.30 | - | - | 0.41 | 0.21, 0.79 |
| 25-44 | - | - | 3.02 | 1.99, 4.58 | - | - | 1.48 | 0.89, 2.45 |
| 45-64 | - | - | ref | - | - | - | ref | - |
| 65+ | - | - | 0.27 | 0.19, 0.40 | - | - | 0.52 | 0.34, 0.80 |
| **Minority ethnicity status** | | | | | | | | |
| White British | - | - | ref | - | - | - | ref | - |
| Ethnic minority | - | - | 1.10 | 0.59, 2.04 | - | - | 1.53 | 0.68, 3.42 |

**Supplementary Table 27. Sensitivity Analyses - Unadjusted odds ratio (OR) and adjusted odds ratio (aOR) for the association between migration status and developing PCC in males during the Omicron period by infection status.**

| **Omicron**  **Sex = Male** | **Regardless of any previous SARS-CoV-2 infections**  **(n = 3,276)** | | | | **With a history of SARS-CoV-2 infection**  **(n = 797)** | | | |
| --- | --- | --- | --- | --- | --- | --- | --- | --- |
| **Variable** | **OR** | **95 CI** | **aOR** | **95 CI** | **OR** | **95 CI** | **aOR** | **95 CI** |
| **Migration status** | | | | | | | | |
| UK born | ref | - | ref | - | ref | - | ref | - |
| Non UK born | 0.58 | 0.14, 2.38 | 0.65 | 0.12, 3.48 | 0.56 | 0.13, 2.36 | 0.75 | 0.13, 4.29 |
| Missing | - * | - * | - * | - * | - * | - * | - * | - * |
| **Age group** | | | | | | | | |
| 0-24 | - | - | 1.75 | 0.69, 4.41 | - | - | 1.24 | 0.47, 3.26 |
| 25-44 | - | - | 0.70 | 0.16, 3.01 | - | - | 0.51 | 0.11, 2.26 |
| 45-64 | - | - | ref | - | - | - | ref | - |
| 65+ | - | - | 0.63 | 0.36, 1.10 | - | - | 0.65 | 0.37, 1.17 |
| **Minority ethnicity status** | | | | | | | | |
| White British | - | - | ref | - | - | - | ref | - |
| Ethnic minority | - | - | 0.82 | 0.20, 3.33 | - | - | 0.67 | 0.16, 2.87 |

**Supplementary Table 28. Sensitivity Analyses - Unadjusted odds ratio (OR) and adjusted odds ratio (aOR) for the association between migration status and developing PCC in females during the Omicron period by infection status.**

| **Omicron**  **Sex = Female** | **Regardless of any previous SARS-CoV-2 infections**  **(n = 4,266)** | | | | **With a history of SARS-CoV-2 infection**  **(n = 1,039)** | | | |
| --- | --- | --- | --- | --- | --- | --- | --- | --- |
| **Variable** | **OR** | **95 CI** | **aOR** | **95 CI** | **OR** | **95 CI** | **aOR** | **95 CI** |
| **Migration status** | | | | | | | | |
| UK born | ref | - | ref | - | ref | - | ref | - |
| Non UK born | 1.34 | 0.69, 2.59 | 0.99 | 0.42, 2.31 | 1.64 | 0.81, 3.34 | 1.22 | 0.48, 3.09 |
| Missing | 1.00 | 0.14, 7.35 | 0.59 | 0.07, 5.07 | 0.73 | 0.09, 5.72 | 0.61 | 0.07, 5.76 |
| **Age group** | | | | | | | | |
| 0-24 | - | - | 0.21 | 0.03, 1.50 | - | - | 0.11 | 0.02, 0.84 |
| 25-44 | - | - | 1.84 | 1.00, 3.41 | - | - | 0.91 | 0.47, 1.75 |
| 45-64 | - | - | ref | - | - | - | ref | - |
| 65+ | - | - | 0.55 | 0.37, 0.82 | - | - | 0.68 | 0.45, 1.03 |
| **Minority ethnicity status** | | | | | | | | |
| White British | - | - | ref | - | - | - | ref | - |
| Ethnic minority | - | - | 1.23 | 0.55, 2.72 | - | - | 1.44 | 0.60, 3.43 |

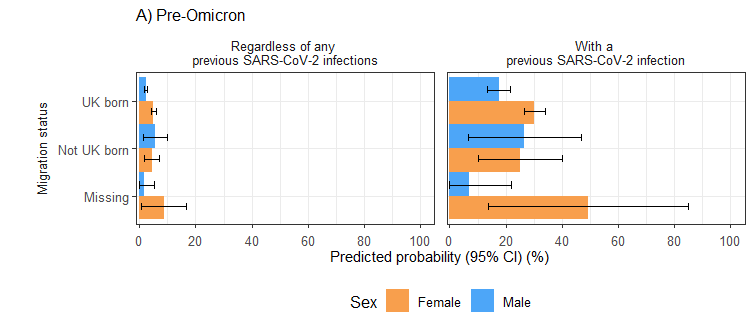

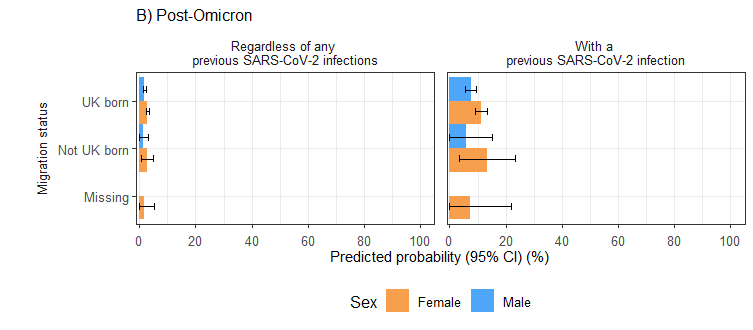

**Supplementary Figure 4. Sensitivity analysis - predicted probabilities of PCC by migration status in the full cohort and individuals with a previous SARS-CoV-2 infection, categorised by reported sex-at-birth during A) pre-Omicron and B) Omicron periods.**

**Supplementary Table 29. Adjusted predicted probability and 95% confidence interval of developing PCC by ‘ethnicity-migration status’ composite variable in males and females during the Pre-Omicron period by infection status.**

| **Pre-Omicron** | **Regardless of any previous SARS-CoV-2 infections** | | **With a history of SARS-CoV-2 infection** | |
| --- | --- | --- | --- | --- |
| **Sex-at-birth** | **Male** | **Female** | **Male** | **Female** |
| **Ethnicity-Migration status** | **(n = 2,883)** | **(n = 3,878)** | **(n = 404)** | **(n = 651)** |
| White British-UK born | 1.93 (1.37, 2.50) | 4.24 (3.54, 4.94) | 13.67 (9.94, 17.39) | 26.05 (22.27, 29.84) |
| White British-Non-UK born | 3.79 (0.00, 8.86) | 2.21 (0.00, 5.22) | 20.37 (0.00, 45.47) | 16.41 (0.00, 37.1) |
| White British-Missing migration status | 5.56 (2.76, 8.36) | 15.32 (10.92, 19.73) | 37.89 (22.36, 53.43) | 53.34 (42.08, 64.6) |
| Ethnic minority-UK born | 1.82 (0.00, 4.34) | 4.27 (1.17, 7.37) | 28.78 (0.00, 62.28) | 34.91 (14.03, 55.8) |
| Ethnic minority-Non-UK born | 7.09 (2.82, 11.36) | 5.64 (2.7, 8.57) | 49.74 (26.37, 73.11) | 34.38 (19.17, 49.58) |
| Ethnic minority-Missing migration status | 1.99 (0.00, 5.89) | 9.43 (1.98, 16.88) | 15.57 (0.00, 44.33) | 58.86 (28.25, 89.46) |

**Supplementary Table 30. Adjusted predicted probability and 95% confidence interval of developing PCC by ‘ethnicity-migration status’ composite variable in males and females during the Omicron period by infection status.** No PP estimates are available for males in the ‘ethnic minority-missing migration status’ group from the Omicron period due to the small sample size.

| **Omicron** | **Regardless of any previous SARS-CoV-2 infections** | | **With a history of SARS-CoV-2 infection** | |
| --- | --- | --- | --- | --- |
| **Sex-at-birth** | **Male** | **Female** | **Male** | **Female** |
| **Ethnicity-Migration status** | **(n = 3,276)** | **(n = 4,266)** | **(n = 797)** | **(n = 1,039)** |
| White British-UK born | 1.42 (0.97, 1.87) | 2.00 (1.53, 2.46) | 5.98 (4.13, 7.82) | 8.22 (6.38, 10.06) |
| White British-Non-UK born | 1.77 (0.00, 5.21) | 1.95 (0.00, 4.62) | 9.42 (0.00, 27.08) | 10.18 (0.00, 23.49) |
| White British-Missing migration status | 6.04 (3.25, 8.84) | 13.46 (9.23, 17.69) | 22.61 (12.98, 32.25) | 37.95 (27.93, 47.97) |
| Ethnic minority-UK born | 1.96 (0.00, 4.7) | 2.82 (0.08, 5.55) | 7.04 (0.00, 16.67) | 13.39 (1.15, 25.64) |
| Ethnic minority-Non-UK born | 0.77 (0.00, 2.28) | 3.64 (1.10, 6.18) | 3.00 (0.00, 8.88) | 19.35 (6.89, 31.8) |
| Ethnic minority-Missing migration status | - | 2.04 (0.00, 6.05) | **-** | 10.23 (0.00, 29.28) |

**Supplementary Table 31. Unadjusted odds ratio (OR) and adjusted odds ratio (aOR) for the association between the ‘ethnicity-migration status’ composite variable and developing PCC in males during the Pre-Omicron period by infection status.**

| **Pre-Omicron**  **Sex = Male** | **Regardless of any previous SARS-CoV-2 infections**  **(n = 2,883)** | | | | **With a history of SARS-CoV-2 infection**  **(n = 404)** | | | |
| --- | --- | --- | --- | --- | --- | --- | --- | --- |
| **Variable** | **OR** | **95 CI** | **aOR** | **95 CI** | **OR** | **95 CI** | **aOR** | **95 CI** |
| **Ethnicity-Migration status** | | | | | | | | |
| White British-UK born | ref | - | ref | - | ref | - | ref | - |
| White British-Non-UK born | 1.98 | 0.47, 8.36 | 2.03 | 0.47, 8.74 | 1.86 | 0.37, 9.23 | 1.64 | 0.32, 8.32 |
| White British-Missing migration status | 3.36 | 1.82, 6.23 | 3.08 | 1.63, 5.79 | 4.33 | 2.05, 9.15 | 4.05 | 1.89, 8.68 |
| Ethnic minority-UK born | 1.72 | 0.41, 7.26 | 0.94 | 0.22, 4.05 | 2.60 | 0.49, 13.82 | 2.63 | 0.46, 14.93 |
| Ethnic minority-Non-UK born | 5.28 | 2.58, 10.8 | 4.04 | 1.91, 8.52 | 8.12 | 3.04, 21.7 | 6.74 | 2.39, 19.04 |
| Ethnic minority-Missing migration status | 2.22 | 0.29, 16.8 | 1.03 | 0.13, 8.05 | 1.62 | 0.18, 14.87 | 1.17 | 0.12, 11.3 |
| **Age group** | | | | | | | | |
| 0-24 | - | - | 1.76 | 0.87, 3.59 | - | - | 0.78 | 0.35, 1.71 |
| 25-44 | - | - | 1.82 | 0.88, 3.76 | - | - | 1.06 | 0.43, 2.62 |
| 45-64 | - | - | ref | - | - | - | ref | - |
| 65+ | - | - | 0.18 | 0.09, 0.34 | - | - | 0.35 | 0.17, 0.71 |

**Supplementary Table 32. Unadjusted odds ratio (OR) and adjusted odds ratio (aOR) for the association between the ‘ethnicity-migration status’ composite variable and developing PCC in males during the Pre-Omicron period by infection status.**

| **Pre-Omicron**  **Sex = Female** | **Regardless of any previous SARS-CoV-2 infections**  **(n = 2,883)** | | | | **With a history of SARS-CoV-2 infection**  **(n = 404)** | | | |
| --- | --- | --- | --- | --- | --- | --- | --- | --- |
| **Variable** | **OR** | **95 CI** | **aOR** | **95 CI** | **OR** | **95 CI** | **aOR** | **95 CI** |
| **Ethnicity-Migration status** | | | | | | | | |
| White British-UK born | ref | - | ref | - | ref | - | ref | - |
| White British-Non-UK born | 0.49 | 0.12, 2.01 | 0.50 | 0.12, 2.08 | 0.63 | 0.14, 2.97 | 0.55 | 0.11, 2.61 |
| White British-Missing migration status | 4.34 | 2.94, 6.43 | 4.40 | 2.92, 6.64 | 2.85 | 1.74, 4.69 | 3.43 | 2.03, 5.8 |
| Ethnic minority-UK born | 1.60 | 0.73, 3.52 | 1.01 | 0.45, 2.25 | 1.43 | 0.56, 3.61 | 1.55 | 0.58, 4.15 |
| Ethnic minority-Non-UK born | 2.21 | 1.24, 3.93 | 1.36 | 0.75, 2.49 | 2.00 | 0.98, 4.07 | 1.51 | 0.73, 3.15 |
| Ethnic minority-Missing migration status | 4.93 | 2.01, 12.08 | 2.44 | 0.95, 6.24 | 4.28 | 1.19, 15.41 | 4.34 | 1.13, 16.74 |
| **Age group** | | | | | | | | |
| 0-24 | - | - | 0.97 | 0.52, 1.81 | - | - | 0.32 | 0.16, 0.64 |
| 25-44 | - | - | 3.05 | 1.99, 4.66 | - | - | 1.51 | 0.9, 2.52 |
| 45-64 | - | - | ref | - | - | - | ref | - |
| 65+ | - | - | 0.27 | 0.19, 0.39 | - | - | 0.52 | 0.33, 0.8 |

**Supplementary Table 33. Unadjusted odds ratio (OR) and adjusted odds ratio (aOR) for the association between the ‘ethnicity-migration status’ composite variable and developing PCC in males during the Omicron period by infection status.** No OR and aOR estimates available for males in the ‘ethnic minority-missing migration status’ group from the Omicron period due to small sample size.

| **Omicron**  **Sex = Male** | **Regardless of any previous SARS-CoV-2 infections**  **(n = 2,883)** | | | | **With a history of SARS-CoV-2 infection**  **(n = 404)** | | | |
| --- | --- | --- | --- | --- | --- | --- | --- | --- |
| **Variable** | **OR** | **95 CI** | **aOR** | **95 CI** | **OR** | **95 CI** | **aOR** | **95 CI** |
| **Ethnicity-Migration status** | | | | | | | | |
| White British-UK born | ref | - | ref | - | ref | - | ref | - |
| White British-Non-UK born | 1.25 | 0.17, 9.28 | 1.25 | 0.17, 9.29 | 1.76 | 0.22, 14.28 | 1.64 | 0.20, 13.38 |
| White British-Missing migration status | 4.66 | 2.59, 8.37 | 4.47 | 2.48, 8.07 | 4.90 | 2.60, 9.26 | 4.62 | 2.43, 8.79 |
| Ethnic minority-UK born | 1.73 | 0.41, 7.30 | 1.39 | 0.32, 6.01 | 1.32 | 0.30, 5.80 | 1.19 | 0.26, 5.42 |
| Ethnic minority-Non-UK born | 0.55 | 0.08, 4.05 | 0.54 | 0.07, 3.99 | 0.47 | 0.06, 3.50 | 0.49 | 0.06, 3.77 |
| Ethnic minority-Missing migration status | - | - | - | - | - | - | - | - |
| **Age group** | | | | | | | | |
| 0-24 | - | - | 1.46 | 0.57, 3.72 | - | - | 1.08 | 0.4, 2.93 |
| 25-44 | - | - | 0.67 | 0.15, 2.91 | - | - | 0.61 | 0.13, 2.8 |
| 45-64 | - | - | ref | - | - | - | ref | - |
| 65+ | - | - | 0.64 | 0.37, 1.12 | - | - | 0.74 | 0.41, 1.34 |

**Supplementary Table 34. Unadjusted odds ratio (OR) and adjusted odds ratio (aOR) for the association between the ‘ethnicity-migration status’ composite variable and developing PCC in males during the Omicron period by infection status.**

| **Omicron**  **Sex = Female** | **Regardless of any previous SARS-CoV-2 infections**  **(n = 2,883)** | | | | **With a history of SARS-CoV-2 infection**  **(n = 404)** | | | |
| --- | --- | --- | --- | --- | --- | --- | --- | --- |
| **Variable** | **OR** | **95 CI** | **aOR** | **95 CI** | **OR** | **95 CI** | **aOR** | **95 CI** |
| **Ethnicity-Migration status** | | | | | | | | |
| White British-UK born | ref | - | ref | - | ref | - | ref | - |
| White British-Non-UK born | 0.95 | 0.23, 3.94 | 0.98 | 0.24, 4.05 | 1.30 | 0.29, 5.74 | 1.27 | 0.29, 5.61 |
| White British-Missing migration status | 7.40 | 4.79, 11.44 | 7.80 | 5.02, 12.12 | 6.39 | 3.9, 10.47 | 7.06 | 4.26, 11.70 |
| Ethnic minority-UK born | 1.65 | 0.59, 4.60 | 1.42 | 0.51, 4.01 | 1.42 | 0.49, 4.15 | 1.73 | 0.58, 5.19 |
| Ethnic minority-Non-UK born | 2.45 | 1.16, 5.19 | 1.86 | 0.86, 4.02 | 2.76 | 1.23, 6.22 | 2.71 | 1.16, 6.34 |
| Ethnic minority-Missing migration status | 1.39 | 0.19, 10.27 | 1.02 | 0.14, 7.77 | 1.00 | 0.13, 7.89 | 1.27 | 0.15, 10.5 |
| **Age group** | | | | | | | | |
| 0-24 | - | - | 0.14 | 0.02, 1.07 | - | - | 0.08 | 0.01, 0.61 |
| 25-44 | - | - | 1.77 | 0.94, 3.32 | - | - | 0.82 | 0.42, 1.62 |
| 45-64 | - | - | ref | - | - | - | ref | - |
| 65+ | - | - | 0.54 | 0.36, 0.80 | - | - | 0.67 | 0.43, 1.03 |
